# How do maternal HIV infection and the early nutritional environment influence the development of infants exposed to HIV *in utero*?

**DOI:** 10.1101/19003871

**Authors:** Marina White, Eleanor Duffley, Ute D. Feucht, Theresa Rossouw, Kristin L. Connor

## Abstract

Malnutrition and infectious disease often coexist in socially inequitable contexts. Malnutrition in the perinatal period adversely affects offspring development and lifelong non-communicable disease risk. Less is known about the effects of *infectious* disease exposure during critical windows of development and health, and links between *in utero* HIV-exposure in the absence of neonatal infection, perinatal nutritional environments, and infant development are poorly defined. In a pilot feasibility study at Kalafong Hospital, Pretoria, South Africa, we aimed to better understand relationships between maternal HIV infection and the early nutritional environment of *in utero* HIV exposed uninfected (HEU) infants. We also undertook exploratory analyses to investigate relationships between food insecurity and infant development. Mother-infant dyads were recruited after delivery and followed until 12 weeks postpartum. Household food insecurity, nutrient intakes and dietary diversity scores did not differ between mothers living with or without HIV. Maternal reports of food insecurity were associated with lower maternal nutrient intakes 12 weeks postpartum, and in infants, higher brain-to-body weight ratio at birth and 12 weeks of age, and attainment of fewer large movement and play activities milestones at 12 weeks of age, irrespective of maternal HIV status. Reports of worry about food runout were associated with increased risk of stunting for HEU, but not unexposed, uninfected infants. Our findings suggest that food insecurity, in a vulnerable population, adversely affects maternal nutritional status and infant development. *In utero* exposure to HIV may further perpetuate these effects, which has implications for early child development and lifelong human capital.

## Introduction

Suboptimal nutrition and poor maternal metabolic health during pregnancy and in the postpartum period are known to impact fetal development, particularly of the brain [1,2], and infant health trajectories after birth [3]. In contrast, the effects of fetal exposure to maternal *infectious* diseases during critical windows of development on offspring growth, neurodevelopment, and lifelong health are less understood. HIV infection has profound effects on maternal physiology and pregnancy outcomes, including increased risk of preterm birth, low birth weight, and increased susceptibility to subsequent infections in infants [4,5]. HIV-exposed, infected (HEI) infants show poorer motor, cognitive, language, and behavioural outcomes compared to controls as early as three months of age [6–9]. However, as global access to antiretroviral therapy (ART) for pregnant and breastfeeding women increases and mother-to-child HIV transmission declines [10], the population of HIV-exposed (*in utero* or during breastfeeding) but *uninfected* (HEU) infants continues to rise. Immune dysfunction, inflammation, metabolic abnormalities, and intestinal dysbiosis persist in HIV-infected individuals on ART, including pregnant women [11]. Some evidence suggests that HEU infants may have persistently altered motor and cognitive development [12–14], albeit to a lesser extent than HEI infants. However, the extent to which inflammatory and metabolic derangements in the mother shape development in their HEU infants remains poorly understood.

The developing brain is also vulnerable to the effects of suboptimal maternal nutrition, as the fetal and neonatal brain depend on nutrition supplied by the mother through transplacental transfer and through breastfeeding and other enteral feeds, to support periods of rapid growth during the perinatal period [15]. Malnutrition (under- or overnutrition) during critical periods of brain development alters neuroanatomical development (neuronal cell growth and proliferation, synapse development, and circuit formation), neurochemistry (synthesis of neurotransmitters, neurotransmitter receptors, and re-uptake mechanisms), and neurophysiology (neurometabolic alterations) [1]. The dependence of the developing brain on adequate nutrition during sensitive periods of its growth and development makes especially the developing fetus, and also the neonate, highly vulnerable to the effects of food and nutrition insecurity.

Interactions between exposure to infectious disease (such as HIV) and malnutrition (such as undernutrition and food insecurity), are important to consider, given that malnutrition and infectious diseases often coexist in socially inequitable contexts. Maternal immunosuppression related to HIV infection may be exacerbated by malnutrition [16], and the comorbidity of these exposures for infants *in utero* and during the breastfeeding period may be more detrimental than the occurrence of one of these circumstances alone. South Africa not only faces the greatest HIV burden globally, with 19% of the world’s population who live with HIV residing in South Africa, but also reports comparable rates of malnutrition and food insecurity [17]. Understanding the extent to which *in utero* HIV exposure, without infant HIV infection, influences infant development is a public health priority [18], and due to the high rates of food and nutrition insecurity, it is therefore also critical to understand how the early nutritional environment interacts with infectious exposures to influence developmental trajectories in HEU infants.

We undertook a prospective cohort pilot feasibility study in Pretoria, South Africa, to determine if we could recruit women through our clinics to study the cooccurring effects of HIV exposure and maternal nutrition on growth and neurodevelopment in infants in early life, and to advance our knowledge on these exposure-outcome relationships. We hypothesised that mothers living with HIV would experience greater household food insecurity, and that maternal HIV infection would influence infant feeding patterns. We also undertook exploratory analyses to investigate overall household food security and diet quality in this cohort, hypothesising that food insecurity alone would be associated with infant feeding patterns, and infant growth and neurodevelopmental outcomes.

## Methods

### Study population

From the obstetric unit at Kalafong hospital, women living with and without HIV were recruited within four days of delivery (HIV infected: n=32, HIV uninfected: n=22) and followed-up at 12 weeks (range 8-16 weeks) postpartum (HIV infected: n=21, HIV uninfected: n=10) at the Maternal and Infant Health Care Strategies Unit (MIHCSU) of the South African Medical Research Council. Study recruitment took place between June-December 2016, with all follow up data collected by March 2017. All infants exposed to HIV were tested for infection at birth and 12 weeks postpartum and were confirmed negative. This study was approved by the Research Ethics Committee of the Faculty of Health Sciences of the University of Pretoria (185-2016) and the Carleton University Research Ethics Board (108870).

### Recruitment and eligibility

A research nurse recruited eligible women after delivery. Exclusion criteria included infant delivery by caesarean section, pregnancy complications (including gestational diabetes mellitus, multiple gestations), or antibiotic exposure during labour or delivery and/or the postpartum period. Women were also ineligible to participate if they were from other regions and would find it difficult to come back for follow-up.

### Data collection

A retrospective medical chart review was conducted after delivery to extract antenatal data, including maternal characteristics (age at conception, ethnicity, parity, gravidity, smoking status, weight during pregnancy); medication use during pregnancy (including antibiotic exposure); illness/infections during pregnancy; and pregnancy outcomes (duration of gestation). At the postpartum follow up visit, mothers completed a questionnaire to assess breastfeeding practices, maternal lifestyle factors (including alcohol intake and smoking), and nutrition (including vitamin supplements, food security, and a 24-hour dietary recall). Where visits to clinics or hospitals occurred between birth and the follow up visit, the patient-retained child health record (Road to Health Chart [19]) was examined to extract data on infant weight, history of illness and medication use.

### Primary outcome measures

Primary outcomes of interest were anthropometry in infants at birth and 12 weeks, weight gain from birth to 12 weeks postpartum, Apgar scores at one and five minutes, and neurodevelopmental status (Guide for Monitoring Child Development [20]) in infants aged 12 weeks.

#### Infant anthropometry

Infant weight, length, and abdominal and head circumference and body mass index (BMI) were measured at birth and in infants aged 12 weeks. Infant anthropometry were age- and sex-standardised using World Health Organization (WHO) growth standards (WHO Anthro software v 3.2.2, January 2011) [21]. A brain weight estimate was calculated using an equation derived by the National Institute of Neurological and Communicative Disorders and Stroke’s Collaborative Perinatal Project [22]:

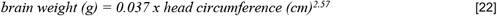

The brain weight estimate was used to calculate the infant brain-to-body weight ratio (BBR)[23]:

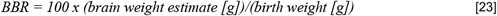

BBR is used as a measure of potential *in utero* asymmetric intrauterine growth restriction (IUGR) and brain sparing [23]. Weight gain from birth to 12 weeks postnatal age (kg/day) was calculated using the weight of an infant at birth and follow up, and the days alive since birth at follow up:

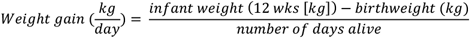

#### Infant neurodevelopment

The Guide for Monitoring Child Development (GMCD) [20] was developed, standardised and validated [24] for use in low- and middle-income countries, including South Africa, to assess expressive and receptive language, play activities, relating and response behaviour, and fine and large movement in infants 1 to 24 months postpartum. An assessment for each infant was carried out once between 8 and 16 weeks postpartum by trained staff at the MIHCSU, which involved the researcher asking the child’s caregiver a series of open-ended questions relating to the child’s development. The GMCD monitors infants who are 1-3 months of age (defined as 1 month to 2 months and 30 days) for achievement milestones listed in the 1-3 month category, and infants who are 3-5 months (3 months+1 day to 4 months+30 days) for achievement of all milestones up to 3-5 months. Infants who were premature (<37 weeks) were assessed according to corrected gestational age. GMCD outcomes were quantified in 2 ways. First, the total number of 1-4 month GMCD milestones attained by each infant, regardless of whether the infant fell was 1-3 or 3-5 months of age at follow up, was quantified for expressive and receptive language, play activities, relating and response behaviour, and fine and large movement. Next, the proportion of infants having attained all milestones in their age category (1-3 months, or 3-5 months) for expressive and receptive language, fine and large movement, play activities and relating/response behaviour, compared to the proportion that had not attained all milestones, was quantified.

### Secondary exposure and outcome measures

#### Maternal food security and dietary recall

A questionnaire was developed to collect maternal reports of food security. Mothers were asked if, in the past 12 months, 1. They and other household members worried that food would run out before they got money to buy more (often true, sometimes true, or never true), 2. The food that they and other household members bought just didn’t last, and there wasn’t any money to get more (often true, sometimes true, or never true), and 3. They and other household members couldn’t afford to eat balanced meals (often true, sometimes true, or never true). Due to our small sample size, maternal reports of ‘often true’ and ‘sometimes true’ were grouped together for analyses as ‘experiences food insecurity’ and compared with ‘never true’ responses.

Maternal dietary recall data collected detailed all food and drink consumed in the day prior to the follow up appointment. Dietary recall data were analysed using FoodFinder3 [25], a dietary analysis software programme developed by the South African Medical Research Council, specific to the nutrient composition of foods in South Africa. The estimated average requirements (EARs) and tolerable upper levels (TULs) for available nutrients from the Institute of Medicine Dietary Reference Intakes were used to evaluate the nutritional adequacy of reported maternal diets [26]. These reference intakes have been used previously to evaluate diet composition in various South African cohorts [27]. A dietary diversity score (DDS) was calculated as an additional measure of diet quality using nine food groups (1. Cereals, roots and tubers, 2. Vegetables and fruits rich in Vitamin A, 3. Other fruit, 4. Other vegetables, 5. Legumes, 6. Meat, poultry and fish, 7. Dairy, 8. Eggs, and 9. Fats and oils) as previously validated and described in South African cohorts [28,29]. Each food group was only counted once.

#### Infant feeding patterns

At follow up, mothers reported whether they were, or had ever, exclusively breastfed their infants. If the infants were currently receiving formula, the mothers provided the age when formula had been introduced. Feeding practices were only available until 8 weeks of age for the youngest infant at follow up, so this was chosen at the cut off for Figure 1 to plot feeding patterns for the whole cohort.

**Figure 1.**
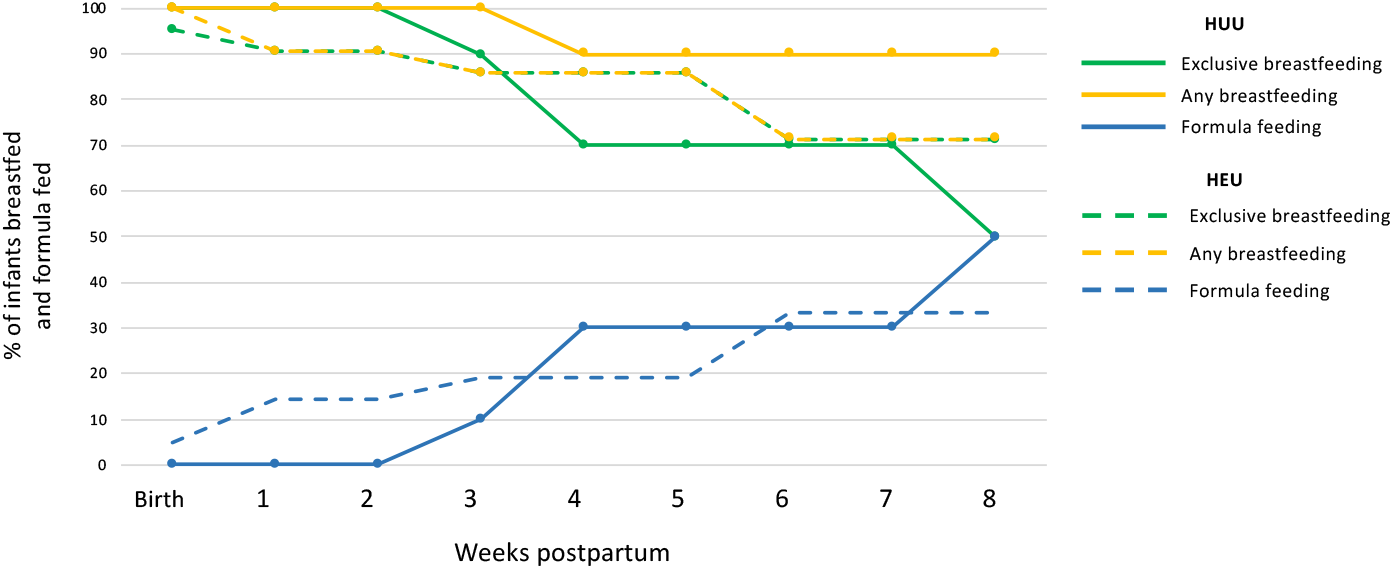
Feeding patterns from birth to 8 weeks postpartum in HUU and HEU infants. There were no differences in the likelihood of being exclusively breastfed (EBF) at 12 weeks of age for HUU (n=10) compared to HEU (n=21) infants ([p>0.05], Fisher’s exact 2-Tail). Data on feeding practices were available for the whole cohort from birth to 8 weeks postpartum. Each point on the line represents the proportion (%) of HUU or HEU exclusively breastfed, receiving any breastmilk, or were formula fed at that time (weeks). HUU = HIV-unexposed, uninfected infant; HEU = HIV-exposed, uninfected infant.

### Data analysis

Data were analysed using JMP 14.0. Data were tested for normality and univariate analysis (ANOVA, Kruskal-Wallis/Wilcoxon test for non-parametric data, or Welch’s test for normal data with unequal variance) was used to compare maternal dietary intake nutrient levels for mothers living with and without HIV. Differences in the probability that mother’s living with compared to without HIV reporting that they experienced food insecurity or had a DDS <4 were assessed using Fisher’s exact test (2-tail).

To explore overall household food security and diet quality in this cohort, and possible relationships between household food insecurity and infant growth, we conducted comparisons of: 1. Anthropometry of infants at birth and 12 weeks of age, 2. Apgar scores (one and five minutes), and 3. GMCD milestones attained in infants aged 12 weeks for infants whose mothers reported on food security using adjusted multiple regression models, and Fisher’s exact test (2-tail) was used to quantify differences for probability of: 1. Stunting at birth or 12 weeks of age, 2. Exclusively breastfeeding at follow up, and 3. Attaining all age-appropriate GMCD milestones, between infants whose mothers reported food insecure conditions compared to those who did not. We also explored whether relationships between suboptimal nutritional environments and infant development would occur to a greater degree in HEU than HUU infants. Using Fisher’s exact test (2-tail), we compared probability of : 1. Stunting at birth or 12 weeks of age, 2. Exclusively breastfeeding at follow up, and 3. Attaining all age-appropriate GMCD milestones for HEU and HUU infant groups.

Maternal data on food security and dietary recall were only available for mothers whose infants attended follow up, when the questionnaires were administered. Therefore, relationships between maternal HIV status, diet, and reports of food (in)security with infant anthropometric and GMCD outcomes at birth and in infants aged 12 weeks were only examined in this subset (HIV infected: n=21, HIV uninfected: n=10) of the original cohort recruited at birth (HIV infected: n= 32, HIV uninfected: n= 22).

Variables that are known or suspected confounders for our outcome measures were included as covariables in regression models where the exposure of interest was maternal reports of food (in)security, and variables with *α*=0.20 were retained through stepwise backward elimination for the final models. Maternal covariables included HIV status, age, education, weight at delivery, gravidity, parity, smoke exposure, breastfeeding practices. Infant included anthropometry, gestational age, sex, age (days) at follow up, weight gain from birth to 12 weeks of age. Variables included in final adjusted models for outcomes at birth and 12 weeks of age can be seen in Supplementary tables S1 and S2. With the pilot study’s small sample size in consideration, we calculated adjusted retrospective power (AdjP) to determine the probability that the any effects of HEU on infant outcomes found during analyses were true with 80% certainty (AdjP>0.80). Findings are presented as unadjusted medians [IQR], with AdjP and p value from ANCOVA.

## Results

### Cohort characteristics

There were no differences among the 31 (HIV infected n=21, HIV uninfected n=10) mother-infant dyads at follow up for maternal weight at delivery, age, level of education, or reports of food insecurity (Supplementary Table S3). No mothers reported consuming alcohol during pregnancy, and all mothers were non-cigarette smokers, however, one woman reported consuming snuff. Of the infants in attendance at follow up, three were born preterm (<37 weeks), including one HUU infant born at 36 weeks, and two HEU infants, born at 35 and 36 weeks (Supplementary Table S3). There were no differences between HUU and HEU infants for infant sex, gestational age or age at follow up at follow up (Supplementary Table S3).

### Food security and nutrients intakes among mothers living with and without HIV

There were no differences between mothers living with and without HIV for probability of reporting household food insecurity (Table 1). Mothers living with HIV had higher intakes of Vitamin D (64.5 [42.0, 84.6] vs. 8.60 [0.38, 20.8], AdjP=0.95, p=0.002) and Se (51.7 [42.1, 73.7] vs. 12.6 [7.41, 34.4], AdjP=0.77, p<.001) compared to mothers living without HIV (Table 1). There were no relationships between maternal HIV status and DDS (Table 1). Full data on absolute nutrient intake levels for mothers living with and without HIV from the 24-hour dietary recall are presented Supplementary table 3.

**Table 1.**
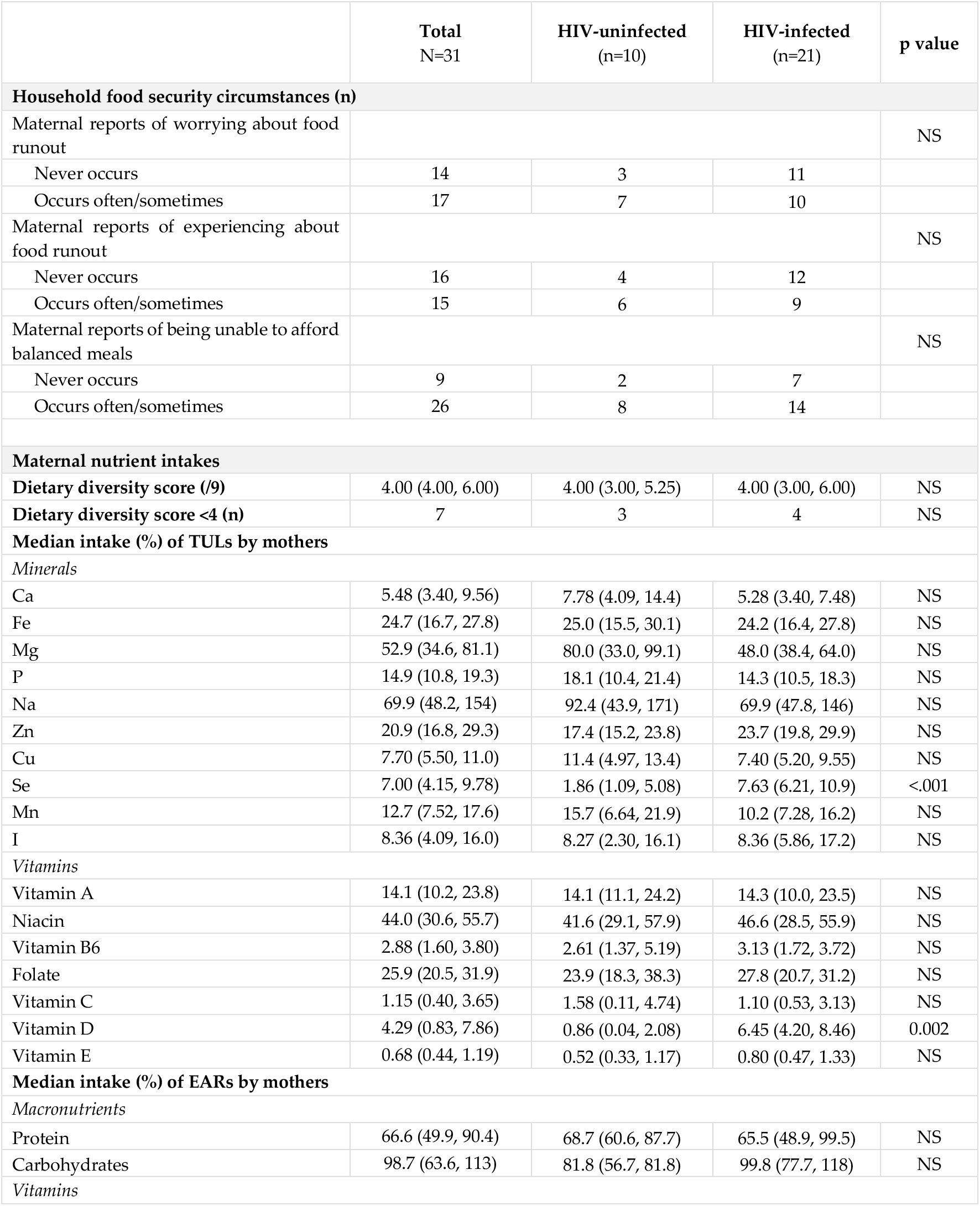
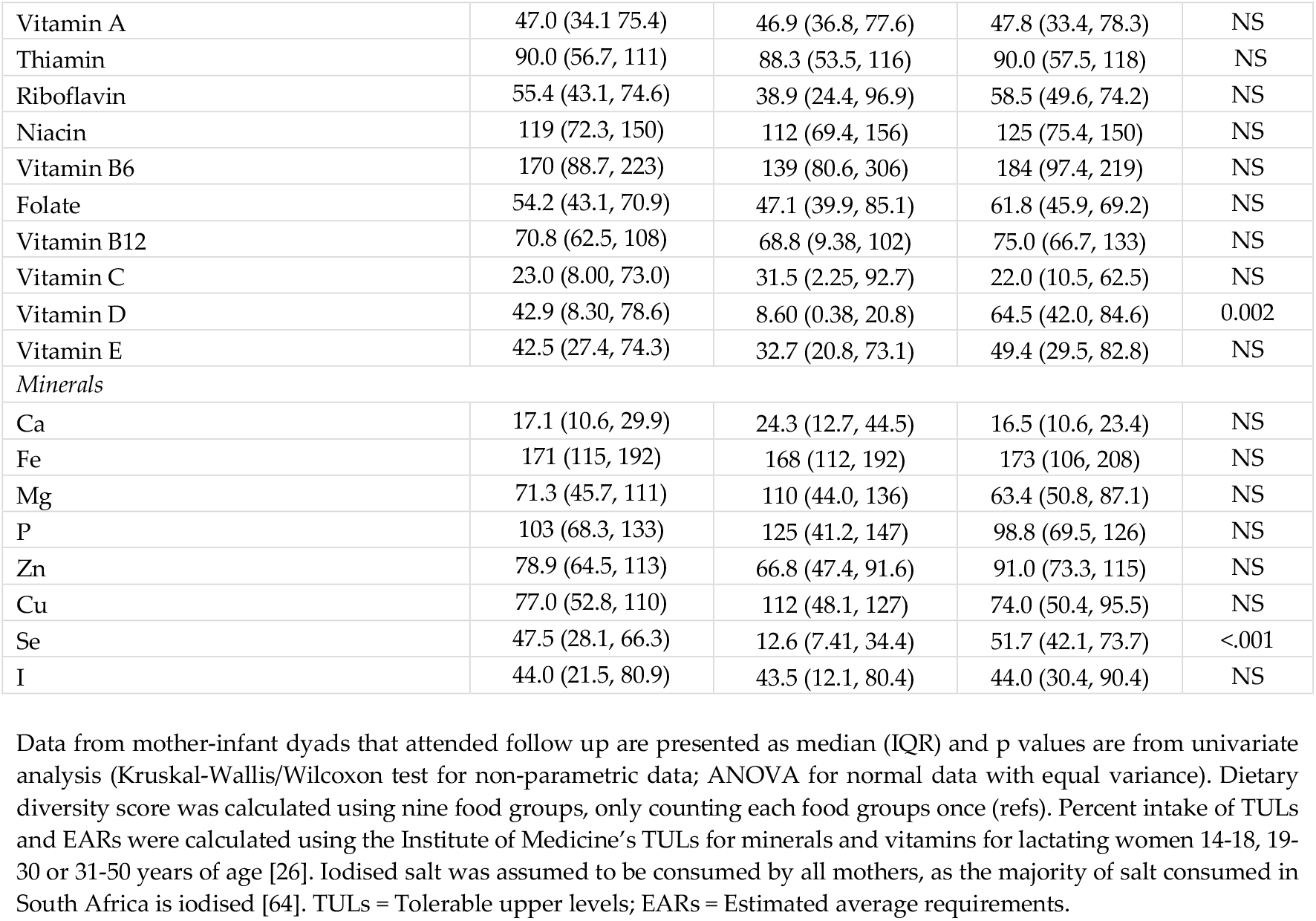
Maternal reports of household food security and nutrient intakes from one 24-hour dietary recall for mothers with and without HIV who attended follow up.

### Relationships between household food insecurity and diet quality

Mothers who worried about or experienced food runout consumed less animal protein compared to those who did not (16.3 [3.85, 30.0] vs. 29.0 [19.3, 39.0], AdjP=0.49, p=0.03 [Table 2]; 15.4 [3.30, 21.4] vs. 29.0 [19.7, 38.0], AdjP=0.68, p=0.01 [Supplementary table S6]). Mothers who experienced food runout consumed less saturated fatty acids (FA)than mothers who did not (6.80 [5.69, 12.9] vs. 9.68 [8.88, 14.7], AdjP=0.07, p=0.03 [Supplementary table S6]). Mothers who worried about food runout consumed lower levels of added sugars (8.00 [0.50, 18.8] vs. 25.6 [12.0, 41.0], AdjP=0.11, p=0.01 [Table 2]) and glucose (1.80 [0.00, 3.55] vs. 3.95 [2.40, 5.00], AdjP=0.19, p=0.04 [Table 2]) compared to those who did not, and mothers who experienced food runout (Supplementary table S6) consumed less fructose (0.40 [0.00, 6.10] vs. 6.50 [1.20, 8.15], AdjP=0.16, p=0.01), sucrose (8.40 [1.00, 17.9] vs. 19.1 [12.4, 30.1], AdjP=0.05, p=0.03), glucose (1.20 [0.00, 3.40] vs. 3.95 [2.75, 5.60], AdjP=0.39, p=0.01), added sugars (8.00 [0.30, 21.0] vs. 17.6 [12.0, 40.5], AdjP=0.05, p=0.04) total sugars (13.1 [7.00, 30.9] vs. 31.2 [24.3, 42.5], AdjP=0.07, p=0.02) and dietary insoluble fibre (1.20 [0.00, 3.20] vs. 3.15 [1.68, 4.08], AdjP=0.18, p=0.04). Overall, very few mothers had intake of vitamins or minerals that was too high, however, TULs were exceeded for magnesium and sodium (Table 2). There were no relationships between reports of household food insecurity and DDS (Table 2, Supplementary table S6).

**Table 2.** Maternal nutrient intake from one 24-hour dietary recall for mothers who report experiencing food insecure compared to those who do not experience food insecurity.

|  | Do you worry about food runout? |  | p value |
| --- | --- | --- | --- |
|  | NO | YES |  |
|  | n=14 | n=17 |  |
| **Dietary diversity score (/9)** | 4.50 (4.00, 6.00) | 4.00 (3.00, 5.50) | NS |
| **Dietary diversity score <4 (n)** | 2 | 5 | NS |
| **Absolute intakes** |  |  |  |
| Macronutrients |  |  |  |
| Moisture (g) | 868 (629, 1029) | 670 (507, 1092) | NS |
| Energy (kJ) | 5601 (4792, 6855) | 5204 (3264, 8103) | NS |
| Nitrogen (g) | 7.15 (5.55, 8.82) | 4.63 (3.93, 6.57) | NS |
| Total protein (g) | 51.4 (43.8, 63.2) | 37.9 (26.3, 53.8) | NS |
| Plant protein (g) | 19.6 (17.1, 26.0) | 22.3 (11.1, 37.5) | NS |
| Animal protein (g) | 29.0 (19.3, 39.0) | 16.3 (3.85, 30.0) | 0.03 |
| Total fat (g) | 36.5 (30.0, 51.8) | 25.8 (20.6, 65.1) | NS |
| Carbohydrates, avail. (g) | 162 (159, 200) | 161 (97.0, 277) | NS |
| Starch (g) | 5.6 (0.18, 10.3) | 0.00 (0.00, 8.1) | NS |
| Glucose (g) | 3.95 (2.40, 5.00) | 1.80 (0.00, 3.55) | 0.04 |
| Fructose (g) | 6.50 (0.90, 7.45) | 0.70 (0.00, 6.30) | NS |
| Sucrose (g) | 17.7 (12.0, 32.9) | 12.0 (1.70, 23.6) | NS |
| Maltose (g)<sup>1</sup> |  | 0.1 |  |
| Galactose<sup>2</sup> |  |  |  |
| Lactose (g) | 0.00 (0.00, 2.10) | 0.00 (0.00, 0.05) | NS |
| Total sugars (g) | 30.3 (23.4, 41.7) | 15.8 (8.05, 34.5) | NS |
| Added sugar (g) | 25.6 (12.0, 41.0) | 8.00 (0.50, 18.8) | 0.01 |
| Total dietary fibre (g) | 13.4 (11.0, 16.7) | 12.2 (6.70, 21.1) | NS |
| Insoluble dietary fibre (g) | 2.75 (1.33, 3.60) | 1.40 (0.00, 3.45) | NS |
| Soluble dietary fibre (g) | 1.85 (1.05, 2.70) | 0.80 (0.00, 3.40) | NS |
| Ash (g) | 6.05 (4.65, 6.53) | 9.30 (4.30, 14.8) | NS |
| Non-starch polysaccharides (g) | 4.20 (2.13, 5.98) | 2.10 (0.00, 6.45) | NS |
| Insoluble NSP (g) | 2.25 (1.15, 3.03) | 1.40 (0.00, 3.05) | NS |
| Soluble NSP (g) | 1.85 (1.05, 2.70) | 0.80 (0.00, 3.40) | NS |
| Lignin (g) | 0.20 (0.10, 0.30) | 0.10 (0.00, 0.30) | NS |
| Amino acids |  |  |  |
| Isoleucine (g) | 1.78 (1.34, 2.26) | 1.24 (0.80, 1.83) | NS |
| Leucine (g) | 3.48 (2.68, 4.06) | 2.18 (1.51, 3.34) | NS |
| lysine (g) | 2.59 (1.84, 3.67) | 1.52 (1.04, 2.65) | NS |
| Methionine (g) | 0.96 (0.88, 1.17) | 0.65 (0.47, 0.91) | 0.01 |
| Phenylalanine (g) | 2.01 (1.53, 2.35) | 1.32 (0.90, 2.11) | NS |
| Threonine (g) | 1.73 (1.33, 2.10) | 1.08 (0.76, 1.71) | NS |
| Tryptophan (g) | 0.61 (0.53, 0.71) | 0.38 (0.30, 0.57) | 0.02 |
| Valine (g) | 212 (1.67, 2.45) | 1.38 (0.98, 2.23) | NS |
| Arginine (g) | 2.53 (1.86, 3.36) | 1.48 (1.18, 2.77) | NS |
| Histidine (g) | 1.14 (0.85, 1.37) | 0.73 (0.44, 1.27) | 0.05 |
| Cystine (g) | 0.62 (0.53, 0.87) | 0.48 (0.31, 0.66) | 0.02 |
| Tyrosine (g) | 1.12 (0.87, 1.44) | 0.79 (0.46, 1.06) | NS |
| Alanine (g) | 1.74 (1.28, 2.28) | 1.16 (0.67, 1.65) | 0.04 |
| Aspartic acid (g) | 3.01 (2.24, 3.71) | 1.87 (1.12, 2.92) | NS |
| Glutamic acid (g) | 6.24 (4.72, 8.31) | 4.43 (1.96, 5.83) | NS |
| Glycine (g) | 1.47 (1.03, 2.10) | 1.02 (0.52, 1.47) | NS |
| Proline (g) | 2.00 (1.47, 2.63) | 1.36 (0.56, 1.87) | 0.05 |
| Serine (g) | 1.65 (1.50, 2.14) | 1.20 (0.86, 1.67) | 0.03 |
| Hydroxyproline (g) | 0.10 (0.06, 0.22) | 0.00 (0.00, 0.11) | NS |
| **Fatty acids and cholesterol** |  |  |  |
| Saturated FA (g) | 9.55 (8.56, 15.2) | 7.88 (5.74, 11.4) | NS |
| Mono-unsaturated FA (g) | 12.5 (10.5, 15.8) | 10.9 (7.19, 21.8) | NS |
| Polyunsaturated FA (g) | 8.70 (6.26, 15.3) | 5.46 (3.37, 20.2) | NS |
| Single trans FA (g)<sup>2</sup> |  |  |  |
| Double trans FA (g)<sup>2</sup> |  |  |  |
| Total trans FA (g) | 0.20 (0.16, 0.82) | 0.06 (0.00, 0.35) | NS |
| Cholesterol (mg) | 291 (236, 544) | 117 (33.0, 420) | 0.04 |
| **Median intake (%) of TULs by mothers** |  |  |  |
| **Minerals** |  |  |  |
| Calcium (Ca) | 5.88 (4.56, 9.55) | 4.52 (2.76, 10.5) | NS |
| Iron (Fe) | 25.6 (21.7, 27.1) | 18.2 (12.8, 31.6) | NS |
| Magnesium (Mg) | 52.1 (44.6, 71.4) | 57.1 (30.0, 97.0) | NS |
| Phosphorus (P) | 16.7 (12.9, 19.9) | 12.1 (9.14, 18.9) | NS |
| Sodium (Na) | 67.5 (48.0, 91.7) | 127 (42.5, 210) | NS |
| Zinc (Zn) | 22.3 (20.1, 29.6) | 19.6 (14.7, 27.1) | NS |
| Copper (Cu) | 8.15 (6.35, 10.6) | 6.90 (3.95, 12.1) | NS |
| Selenium (Se) | 7.56 (5.41, 11.6) | 6.18 (1.86, 9.73) | NS |
| Manganese (Mn) | 12.8 (8.46, 18.2) | 12.7 (5.59, 18.9) | NS |
| Iodine (I) | 7.82 (3.73, 12.8) | 13.6 (4.59, 19.1) | NS |
| **Vitamins** |  |  |  |
| Vitamin A | 14.2 (11.2, 18.8) | 14.1 (8.57, 26.9) | NS |
| Niacin | 47.4 (42.0, 55.6) | 40.0 (24.4, 60.6) | NS |
| Vitamin B6 | 3.20 (2.74, 3.68) | 2.14 (1.22, 4.17) | NS |
| Folate | 27.8 (21.3, 29.6) | 24.1 (19.8, 48.6) | NS |
| Vitamin C | 1.28 (0.90, 2.45) | 0.60 (0.08, 6.38) | NS |
| Vitamin D | 4.73 (4.15, 8.76) | 1.39 (0.39, 7.86) | NS |
| Vitamin E | 0.73 (0.49, 1.21) | 0.68 (0.41, 1.30) | NS |

Data from mother-infant dyads that attended follow up are presented as mean ± SD or median (IQR) and p values are from univariate analysis (Kruskal-Wallis/Wilcoxon test for non-parametric data; ANOVA for normal data with equal variance). Dietary diversity score was calculated using nine food groups, only counting each food groups once (refs). Percent intake of TULs were calculated using the Institute of Medicine's TULs for minerals and vitamins for lactating women 14-18, 19-30 or 31-50 years of age [26]. Iodised salt was assumed to be consumed by all mothers, as the majority of salt consumed in South Africa is iodised [64]. TULs = Tolerable upper levels. <sup>1</sup>Only one mother consumed maltose. <sup>2</sup>None consumed by mothers.

Overall, a large proportion of mothers were at risk of inadequate intake of macronutrients, vitamins and minerals (Figure 2). Food insecurity was associated with an increased risk of inadequate intake (median %EAR met) of vitamin B12 amongst mothers who reported experiencing (66.7 [12.5, 91.7] vs. 89.6 [67.7, 144], AdjP=0.20, p=0.01 [Supplementary figure S1]) food runout, or an inability to afford balanced meals (66.7 [19.8, 96.9] vs. 108 [70.8, 150], AdjP=0.05, p=0.04 [Supplementary figure S2]) compared to mothers who did not. Of the mothers who reported worrying about or experiencing food runout, or inability to afford balanced meals, 76.5%, 86.7%, and 81.8% were at risk for inadequate intake, respectively.

**Figure 2.**
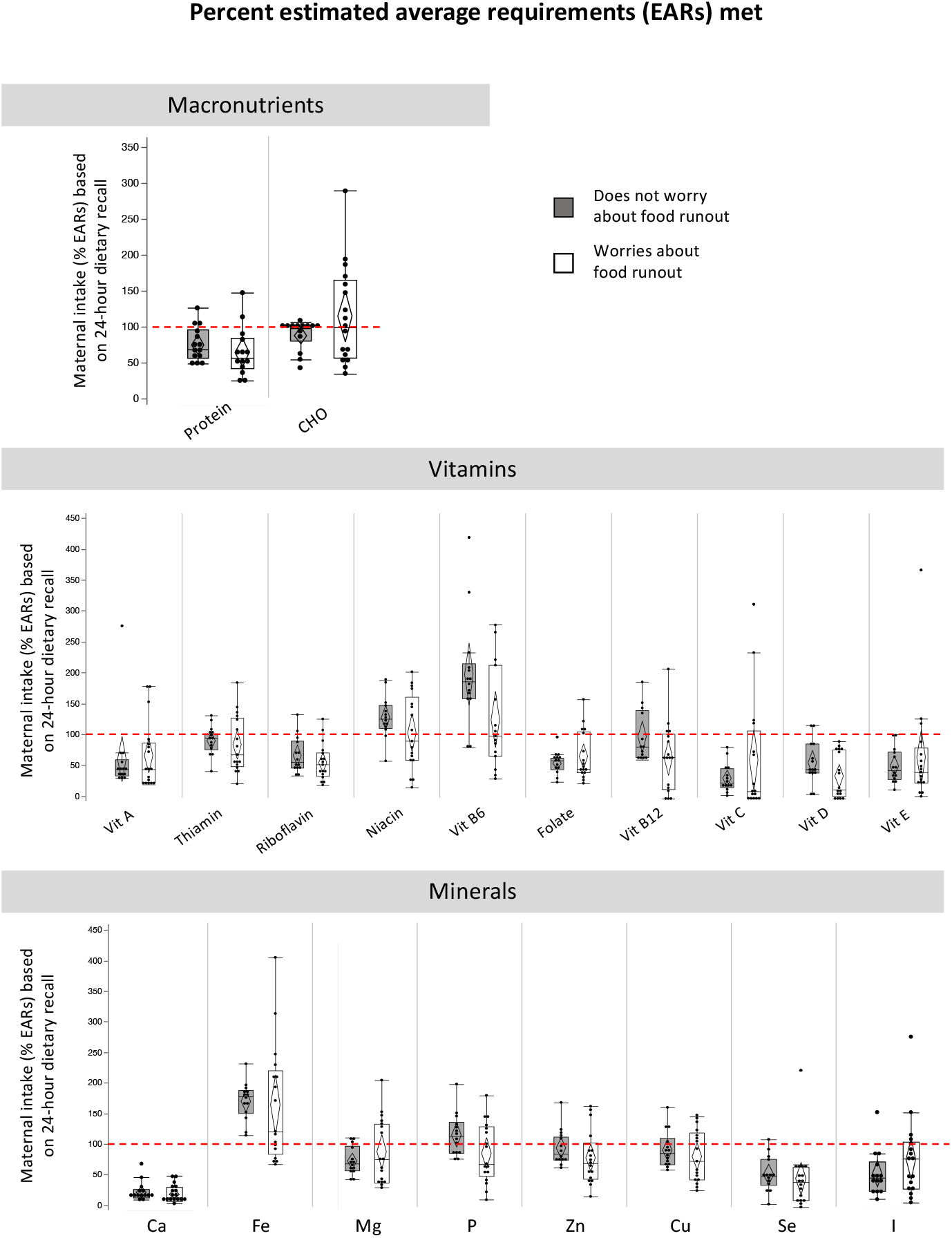
Maternal intake of estimated average requirements for macronutrients, vitamins and minerals for mothers who report worrying about food runout (compared to not worrying). Maternal reports of food insecurity did not associate with intake levels of macronutrients, vitamins or minerals. Many women, irrespective of food security reports, are at risk of inadequate macronutrient, vitamin and mineral intakes. Percent intake of EARs for nutrients were calculated for lactating women 14-18, 19-30 or 31-50 years of age [26]. No EARs are available for total fat. Calculations for EAR for total protein considered maternal weight at time of dietary recall. Data are % intake of EAR reported in maternal dietary recall for macronutrients (quartiles, median lines and 95% confidence diamonds, *p<0.05 [ANOVA for normal distribution/equal variance; Kruskal-Wallis/Wilcoxon test for non-parametric data; or Welch’s test for normal data/unequal variance]). CHO = carbohydrates.

### Influence of food insecurity on infant Apgar scores and growth outcomes at birth

Maternal reports of food insecurity did not influence infant gestational age at birth (Supplementary Table S4). However, infants who were from households where mothers reported worrying about food runout (compared to those who had not worried) had higher BBR at birth (10.4 [9.70, 12.0] vs. 9.65 [8.90, 10.3], AdjP=0.40, p=0.04, Figure 3A). Maternal reports of food insecurity (worry about or experience food runout, or inability to afford balanced meals compared to those who did not experience these circumstances) were not associated with any other infant anthropometry at birth (Supplementary Table S4).

**Figure 3.**
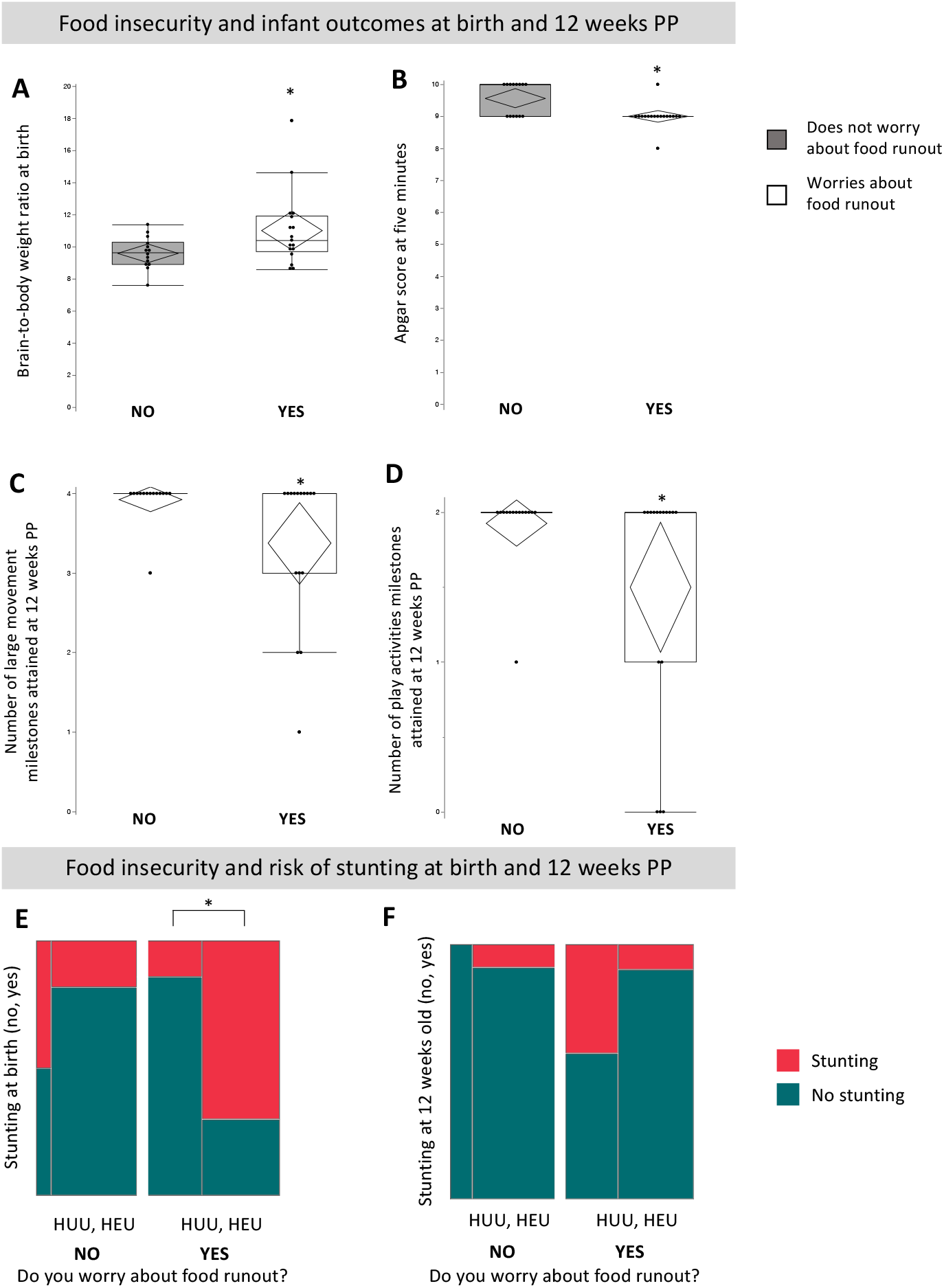
Maternal reports of worrying about food runout affect infant growth and neurodevelopment at birth and 12 weeks of age, and cooccurrence of maternal HIV and food insecurity increases risk of stunting at birth. Maternal reports of worrying about food insecurity (compared to not worrying) associate with higher brain-to-body weight ratio at birth (A; AdjP=0.40, p=0.04), lower Apgar score at five minutes postpartum (B; AdjP=0.55, p=0.001), and attainment of fewer large movement (C; AdjP=0.44, p=0.03) and play activity (D; AdjP=0.58, p=0.02) milestones. Amongst infants whose mothers report worrying about food runout, risk of stunting at birth is greater for HEU compared to HUU infants (E; p=0.04, Fisher’s exact test). The red line represents the proportion of infants who had stunting at birth or 12 weeks PP. Outlier box plots are measured anthropometry, Apgar scores and number of GMCD milestones attained (quartiles, median lines and 95% confidence diamonds, *p<0.05 [ANCOVA]). Mosaic plots are proportion (%) of HUU or HEU infants who have stunting (<-2 SD length-for-age standardised according to WHO child growth standards [21]) at birth and 12 weeks old. GMCD = Guide for monitoring child development. HUU = HIV-unexposed, uninfected infant; HEU = HIV-exposed, uninfected infant.

Apgar scores at one minute were not impacted by maternal reports of food security; however, Apgar scores at five minutes were slightly, but not notably, lower for infants whose mothers reported sometimes or often worrying about food runout (compared to those who never worried; 9.00 [9.00, 9.00] vs. 10.0 [9.00, 10.0], AdjP=0.55, p=0.02 [Figure 3B]). There were no differences in infant Apgar scores at one and five minutes, or trajectory of scores between one and five minutes, for HUU and HEU infants whose mothers reported food insecurity compared those who did not (Supplementary Figure 4).

### Influence of food insecurity on growth outcomes in 12 week old infants

There were no differences for infant head circumference, weight, BMI, or length in infants aged 12 weeks, or weight gain from birth to 12 weeks postpartum in relation to measures of food security (Supplementary Table S4).

### HEU may increase vulnerability to effects of food insecurity on risk of stunting at birth

Among infants whose mothers reported worrying about food runout, HEU infants had increased risk of stunting at birth compared to HUU infants (p=0.04, Fisher’s exact test [2-tail] [Figure 3E]). Maternal HIV status did not influence the relationship between maternal reports of food insecurity and any other infant outcomes at birth or 12 weeks postpartum.

### Influence of food insecurity on neurodevelopmental outcomes in 12 week old infants

All but one infant, who was HUU, received a GMCD assessment at their follow up appointment. Maternal reports of worrying about food runout (compared to not worrying) associated with attainment of fewer large movement (4.00 [3.00, 4.00] vs. 4.00 [4.00, 4.00], AdjP=0.44, p=0.03 [Supplementary Table S4]) and play activities milestones (2.00 [1.00, 2.00] vs. 2.00 [2.00, 2.00], AdjP=0.58, p=0.02 [Supplementary Table S4]). The number of expressive or receptive language, fine movement, or relating and response behaviour milestones attained in infants aged 12 weeks was not affected by maternal reports of food insecurity (Supplementary Table S4).

The probability of attaining all 1-3 month GMCD expressive and receptive language, large movement, play activities and relating and response behaviour milestones did not associate with maternal reports of household food insecurity (Supplementary figure S5A, C, E). No comparisons were made for fine movement outcomes between infants exposed to food insecure conditions compare to those who were not, as all infants who were 1-3 months of age at follow up met all age-appropriate fine movement milestones. At 3-5 months, there were no differences in the proportion of infants who attained fine movement and relating and response behaviour milestones based on maternal reports of food security (Supplementary figure S5B, D, F). No comparisons were made for expressive and receptive language, large movement, or play activities, as all infants 3-5 months of age at follow up had met these milestones. It was not possible to further stratify these comparisons based on maternal HIV status due to our small sample size.

## Discussion

As infectious disease and malnutrition often coexist, it is critical to understand how food insecure circumstances and maternal malnutrition influence the developmental trajectories of the increasing number of children born each year who are HIV-exposed, but uninfected. Amongst children infected with HIV, the cooccurrence of malnutrition is known to exacerbate the disease’s adverse effects, and associates with poorer prognosis [30]. HIV-infection has a definite impact on growth and neurodevelopmental outcomes of children [6–9], and malnutrition, which alone positions children to be more vulnerable to more frequent and severe infections [31], may have an additive negative effect on growth and health trajectories [32].

In this small pilot feasibility study, exploratory analyses found that HEU infants exposed to household food insecurity had increased risk of stunting at birth compared to HUU infants whose mothers also reported food insecure conditions, suggesting that HEU infants may be more vulnerable to the programming effects of suboptimal nutrition *in utero* and postnatally than their HUU counterparts. Stunting is the most common manifestation of infant undernutrition globally [33], increasing morbidity and mortality in children in early life, and increasing one’s risk of metabolic disease while reducing neurodevelopmental and economic capacity later in life [34]. Stunting is common amongst children living with HIV who are under 5 years and improves overtime with ART treatment [35], however, the extent to which HEU children are persistently at risk for stunting is not well understood. In our cohort, there were 11 infants who were stunted at birth, and five who were stunted at 12 weeks postpartum. When examining stunting by HIV exposure groups, we observed that amongst infants whose mothers worried about food runout, HEU infants were at an increased risk of stunting compared to HUU infants at birth, but not at follow up. This finding suggests that the effects of *in utero* HIV exposure and food insecurity on infant length at birth may not persist at 12 weeks postpartum, and infants with low length-for-age at birth may catch up in growth in the first few months of age. Of note, the number of HUU infants 12 weeks of age in our cohort was small, and as such, comparisons between groups should be interpreted with caution. Our findings should be replicated in larger cohorts to determine whether HEU infants exposed to food insecure conditions are indeed more vulnerable to stunting than their HUU counterparts, and whether or not these infants may exhibit catch up growth, an effect known to associate with poor metabolic and cardiovascular outcomes later in life [36].

Suboptimal maternal nutrition during the prenatal period is also known to associate with asymmetric IUGR and brain sparing [23], resulting in a higher BBR at birth and increasing a child’s risk of motor hyperactivity, and poorer language, cognitive, and motor outcomes at 10 years of age [37], stunted immune functioning [38,39] and increased risk of metabolic disease later in life [40]. We found higher BBR at birth amongst infants whose mothers reported food insecure conditions, consistent with reports of associations between maternal malnutrition and brain sparing [23]. Maternal HIV infection is also implicated as a risk factor for IUGR [41], however, risk of asymmetric IUGR and brain sparing for infants exposed to HIV *in utero* is not well characterised, and we found no differences in BBR between HUU and HEU infants exposed to food insecure conditions versus not. This suggests that HEU infants in this cohort were not at an increased risk of brain sparing. To our knowledge, this is the first study to use a measure of BBR to look at differential risk of brain sparing among HUU and HEU infants exposed to suboptimal maternal nutrition *in utero* and postnatally, and larger cohort studies with higher power should further investigate these relationships.

The detrimental impacts of maternal malnutrition on infant neurodevelopment are well established [42], and whilst children perinatally infected with HIV have lower Apgar scores [43], and show poorer motor, cognitive and behavioural development compared to uninfected counterparts as early as 3 months of age [6–9], the neurodevelopmental trajectories of HEU children are poorly understood in part because studies often vary greatly in design, measurement tools, cohort demographics, and findings [13,14,44–47]. Here, we observed slightly lower Apgar scores at five minutes amongst infants whose mothers reported worrying about or experiencing food runout, irrespective whether or not the infant was HUU or HEU. Importantly, these differences were small with low power, and their clinical relevance is unlikely. We also found associations between maternal reports of food insecurity and attainment of play activities and large movement milestones in infants aged 12 weeks, irrespective of maternal HIV status. Further, both HUU and HEU infants 1-3 months of age whose mothers reported food insecure conditions did not meet GMCD 1-3 months milestones for receptive language, large movement, relating and response behaviour or play activities. This is in contrast to HUU and HEU infants aged 3-5 months who met all 3-5 month GMCD neurodevelopmental milestones (except for fine movement), irrespective of household food security status. In disagreement with our hypothesis, these findings suggest that the effects of household food insecurity, possibly resulting in maternal malnutrition, on neurodevelopment were not exacerbated by exposure to HIV in the womb. These data also suggest that whilst food insecure circumstances influence neurodevelopmental milestone attainment in infants who are 1-3 months of age, effects of food insecurity on GMCD milestones may not persist beyond 2 months. As the absence of effect at 3-5 months may be owing to our small sample size and low power, further study in a larger cohort and more longitudinal data are required to determine how infant development may change overtime in response to household food insecurity.

Maternal nutrient intakes during pregnancy and the postpartum period are critical to supporting rapid growth and development of the infant [1], and maternal diet in part determines the nutritional composition of breastmilk [48]. Differences in maternal nutrient intakes between mothers living with and without HIV were minimal, and mothers were at risk for inadequate macronutrient intakes irrespective of food insecurity reports, and mothers experiencing food insecurity consumed less animal protein, sugars and insoluble fibre, and more saturated FA. We also found maternal reports of experiencing food insecurity associated with lower vitamin B12, and a large proportion of mothers, irrespective of food insecurity circumstances, were at risk for inadequate intakes. Inadequate maternal vitamin B12 intakes have shown to cause secondary deficiencies in breastfed infants in the first six months of life, leading to delayed growth and neurodevelopment [49]. Iron intake was not influenced by food insecurity, and it is promising that most mothers exceeded EARs, but not TULs, for iron, which may be the result of public health efforts to prevent and treat anaemia in pregnant women in South Africa. Early detection of maternal nutrient deficiencies allows early intervention and potential mitigation of adverse pregnancy outcomes and long-term developmental impacts for infants who are breastfed, and these preliminary results suggest that food insecurity may influence maternal nutrient status. Importantly, our single dietary recall prevented us from determining whether these deficiencies may have persisted long term or are small fluctuations, limiting our ability to infer the degree they may have secondary consequences for the mother and developing infant.

As ART coverage and viral suppression for pregnant women living with HIV has increased, recommendations around postnatal feeding practices have evolved to promote breastfeeding for women living with HIV exclusively for the first 6 months postpartum [50], with continuation of breastfeeding through mixed feeds thereafter until 24 months postpartum [51]. Breastfeeding benefits include increased child cognitive scores, decreased risk of contraction of common childhood infections, and decreased rates of overweight and diabetes in later life [52]. We observed slightly higher retention of EBF amongst mothers living without HIV compared to those living with HIV. Food insecurity is a known barrier to exclusive breastfeeding [53], and mothers experiencing food insecurity may be more likely to return to work soon after birth [54], or may have challenges maintaining milk supply due to inadequate nutrition [55]. In agreement with this, we found that mothers who never experienced food runout or were always able to afford balanced meals were more likely to be exclusively breastfeeding at follow up. Our short follow up time and small sample size limited our ability to examine the effect of exclusive breastfeeding duration on infant outcomes. Yet, infant growth and neurodevelopment from birth to 12 postpartum did not appear to be influenced by exclusive breastfeeding status at 12 weeks postpartum.

Further, whilst it is known that altered maternal endocrine and metabolic status can restructure the breast milk microbiome and immune and nutrient factors therein [56,57], how maternal HIV infection may alter breastmilk composition is not well understood. Investigation of breastmilk immune factors among women living with HIV remains limited, and among the few studies that have measured these factors, varied results have been reported. Higher levels of non-specific IgA [58] and IgG [58,59] have been measured in breastmilk from women living with HIV, and higher levels of fucosylated human milk oligosaccharides in breast milk are associated with lower infant mortality among HEU infants, but not HEI, during breastfeeding [60]. There is opportunity to better understand how the benefits of exclusive breastfeeding, including improved maternal health outcomes [61] and constituents in breastmilk [62], could be protective against any adverse effects of exposure to HIV in the womb on child growth and neurodevelopment.

Whilst our study’s small sample size and limited longitudinal data restricts the generalisability of our findings and the power of our analyses, we were successful in recruiting women to study the cooccurring effects of food insecurity and *in utero* HIV exposure on maternal dietary intakes and infant outcomes at two time points, and we are now conducting a larger prospective pregnancy and birth cohort study at Kalafong Hospital where we will further investigate relationships that have emerged in exploratory pilot study analyses. There was loss to follow up in this cohort, which may have resulted in sample bias at follow up. While multiple attempts were made to contact mothers whom missed follow up appointments, this population is highly mobile and study participant retention remains difficult. To our knowledge, this prospective cohort study is amongst the first few investigating relationships between maternal nutrition and food security and infant growth and neurodevelopment among a population of HIV-exposed, uninfected infants in comparison to HIV-unexposed controls.

Overall, our findings suggest that food insecurities, and the likely ensuing poor maternal nutritional status, adversely affect the growth and neurodevelopment of a South African cohort of HUU and HEU infants in the first four months of life, and at least for some measures, the effects of a suboptimal early life nutritional environment may be most detrimental for infants exposed to HIV in the womb. The perinatal window is a key period to target to improve infant developmental outcomes. Whether interventions that focus on maternal/child nutritional status and food security at this time improve outcomes for the 1.5 million HIV-exposed infants born annually, who may be even more vulnerable to the programming effects suboptimal nutrition *in utero* and postnatally [63], remains to be determined.

## Data Availability

The datasets generated during and/or analysed during the current study are available from the corresponding author on reasonable request.

## Acknowledgments

The authors would like to thank the participating families and health care workers in Pretoria, for without whom this research would not be possible, and Dr. Chrisna Durandt, who performed the flow cytometry.

This research is funded by the Collaborative Initiative for Paediatric HIV Education and Research (CIPHER), the Faculty of Science, Carleton University, and the Canadian Institutes of Health Research (CIHR). MW is supported by a Canadian Graduate Scholarship-Master’s from CIHR.

## Author Contributions

Conceptulalisation: KLC, TR, UF; Methodology: KLC, MW, TR; Investigation: MW, KLC, ED, TR, UF; Analysed data: MW, ED, KLC; Wrote paper: MW, KLC; Reviewed and revised paper: TR, UF, ED, MW, KLC.

---

**Supplementary figure S1.**
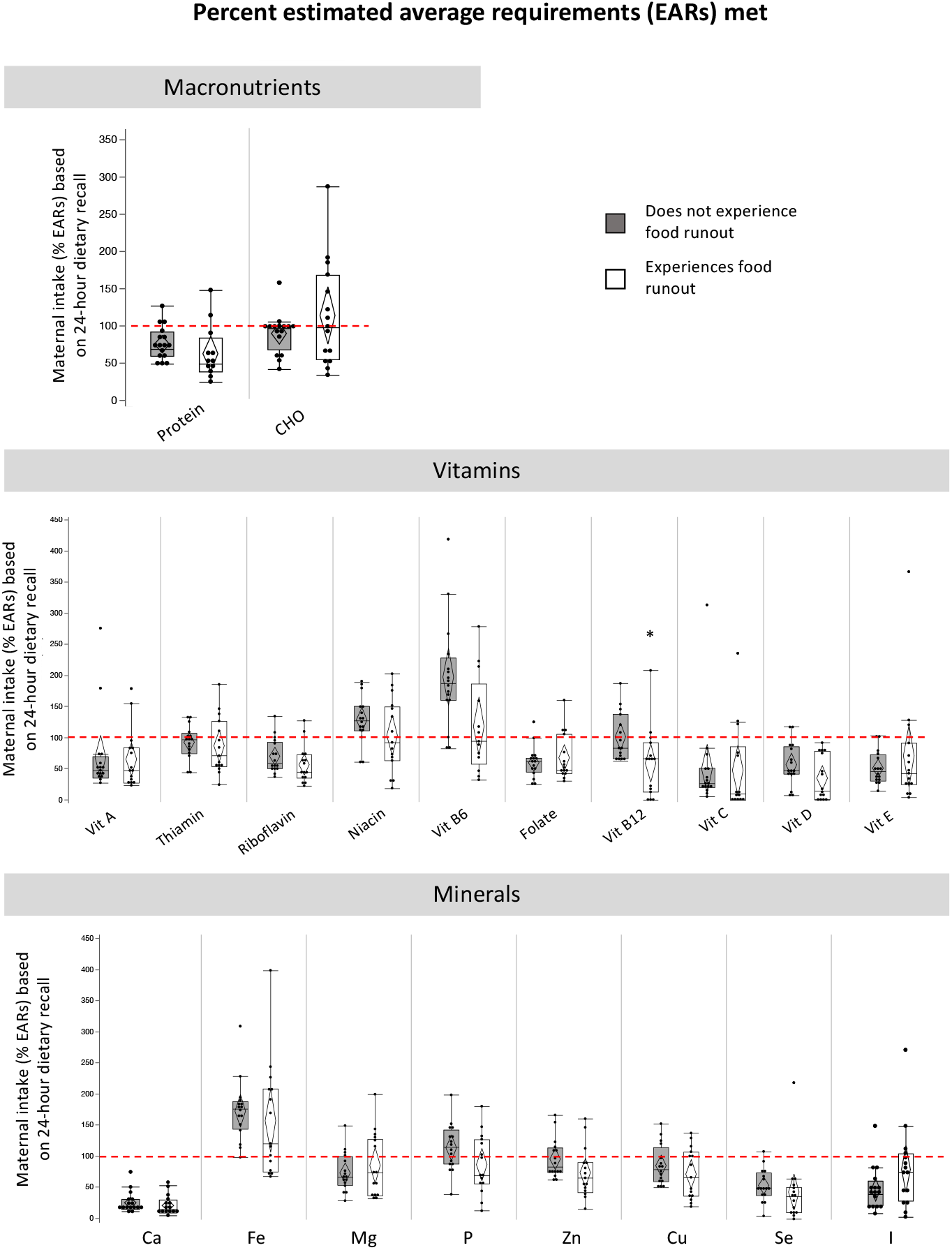
Maternal intake of estimated average requirements for macronutrients, vitamins and minerals for mothers who report experiencing food runout (compared to not). Maternal reports of food insecurity did not associate with intake levels of macronutrients or minerals. Maternal reports of experiencing food runout associated with lower intake of vitamin B12 [B; AdjP=0.20, p=0.01]). Many women, irrespective of food security reports, are at risk of inadequate macronutrient, vitamin and mineral intakes. Percent intake of EARs for nutrients were calculated for lactating women 14-18, 19-30 or 31-50 years of age [26]. No EARs are available for total fat. Calculations for EAR for total protein considered maternal weight at time of dietary recall. Data are % intake of EAR reported in maternal dietary recall for macronutrients (quartiles, median lines and 95% confidence diamonds, *p<0.05 [ANOVA for normal distribution/equal variance; Kruskal-Wallis/Wilcoxon test for non-parametric data; or Welch’s test for normal data/unequal variance]). CHO = carbohydrates.

**Supplementary figure S2.**
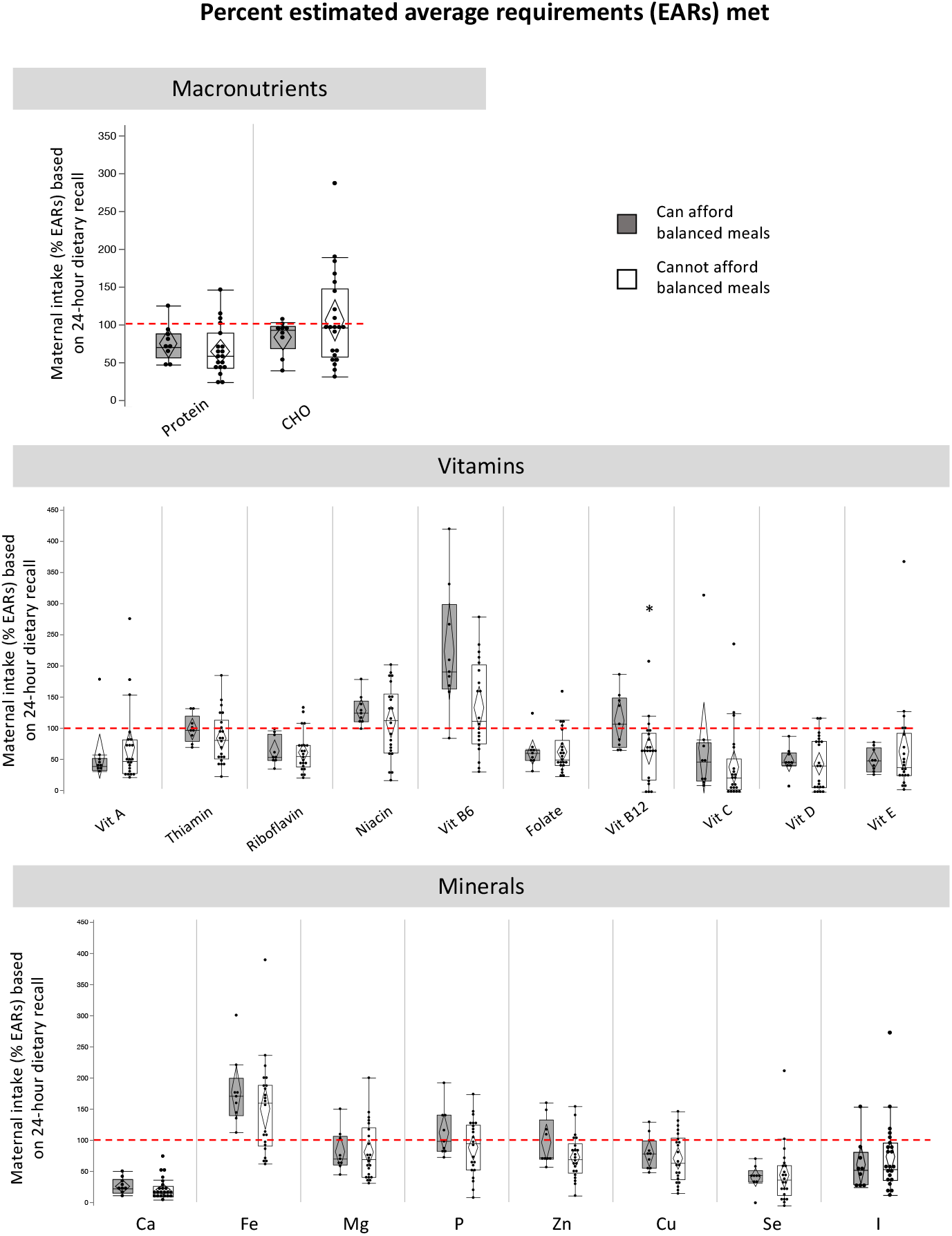
Maternal intake of estimated average requirements for macronutrients, vitamins and minerals for mothers who report inability to afford balanced meals (compared to not). Maternal reports of food insecurity did not associate with intake levels of macronutrients or minerals. Maternal reports of inability to afford balanced meals associated with lower intake of vitamin B12 (C; AdjP=0.05, p=0.04). Many women, irrespective of food security reports, are at risk of inadequate macronutrient, vitamin and mineral intakes. Percent intake of EARs for nutrients were calculated for lactating women 14-18, 19-30 or 31-50 years of age [26]. No EARs are available for total fat. Calculations for EAR for total protein considered maternal weight at time of dietary recall. Data are % intake of EAR reported in maternal dietary recall for macronutrients (quartiles, median lines and 95% confidence diamonds, *p<0.05 [ANOVA for normal distribution/equal variance; Kruskal-Wallis/Wilcoxon test for non-parametric data; or Welch’s test for normal data/unequal variance]). CHO = carbohydrates.

**Supplementary figure S3.**
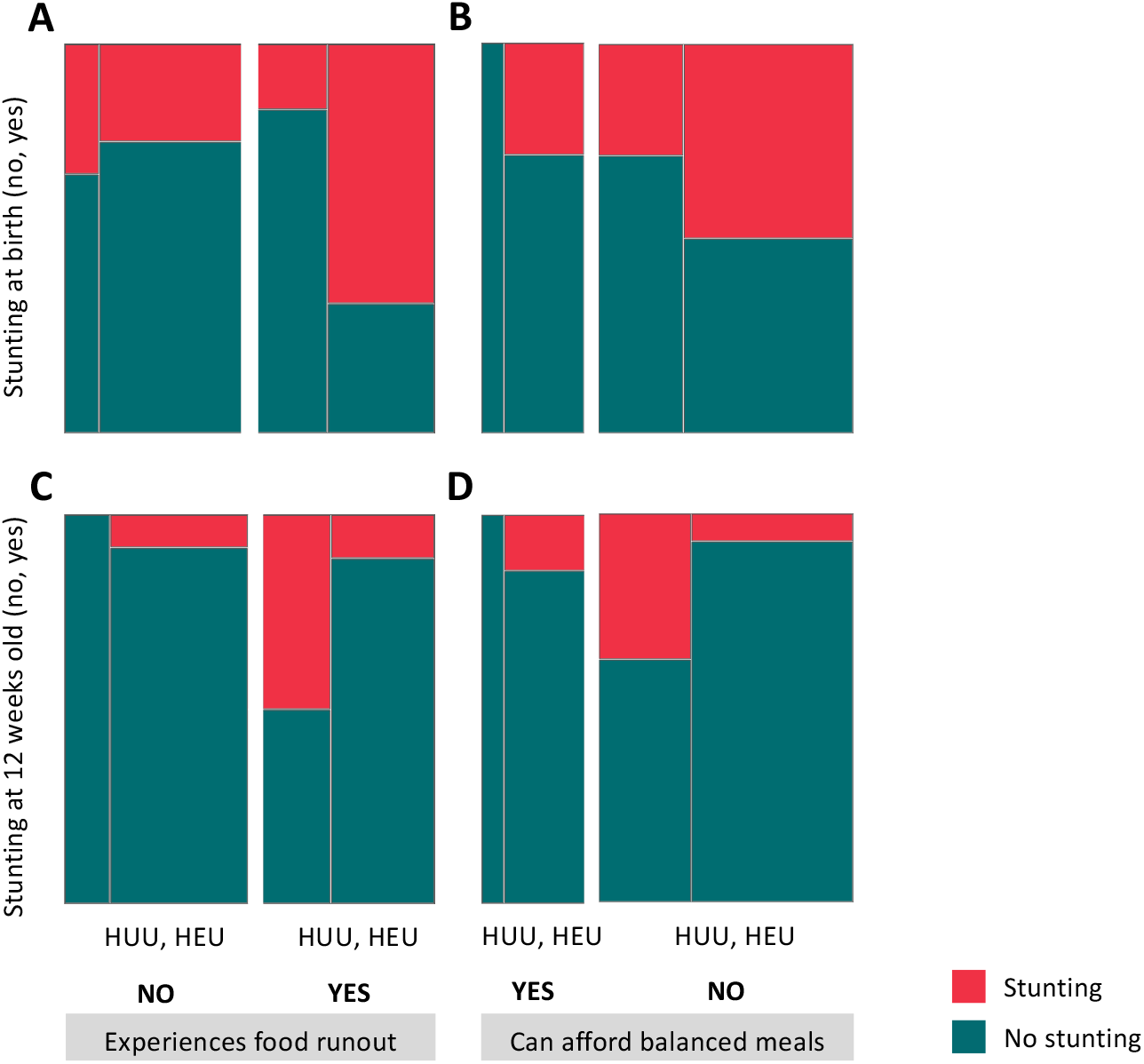
Cooccurrence of maternal HIV and food insecurity does not influence risk of stunting at birth. Maternal reports of experiencing food runout or inability to afford balanced meals (compared to not experiences these circumstances) does not influence risk of stunting at birth for HEU compared to HUU infants (A-D). Mosaic plots are proportion (%) of HUU or HEU infants who have stunting (<-2 SD length-for-age standardised according to WHO child growth standards [21]) at birth and 12 weeks old. GMCD = Guide for monitoring child development. HUU = HIV-unexposed, uninfected infant; HEU = HIV-exposed, uninfected infant.

**Supplementary figure S4.**
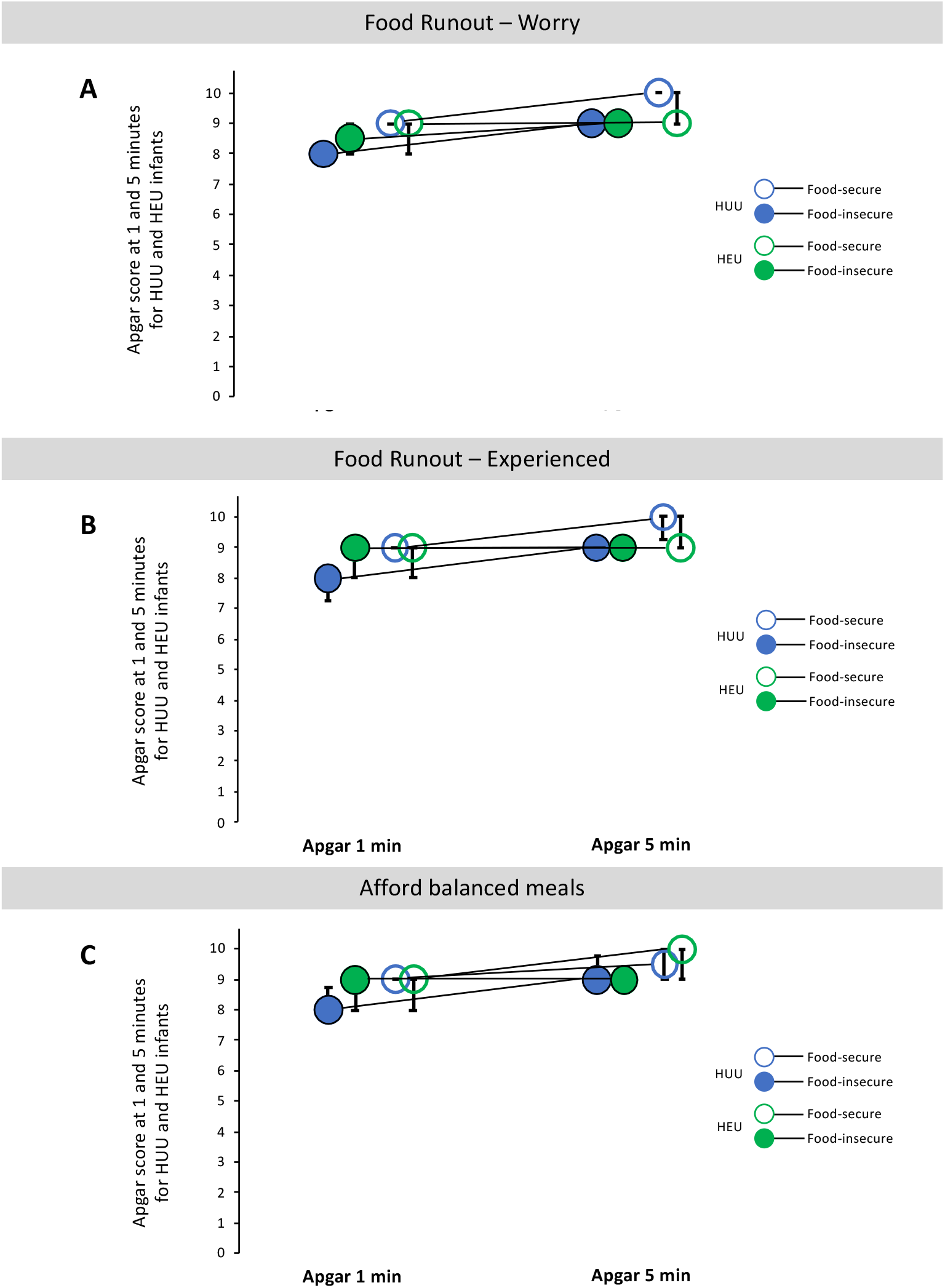
Apgar scores at one and five minutes stratified by HIV exposure status and reports of food insecurity. Apgar scores at one and five minutes after birth were not different between HUU (n=10) and HEU (n=21) infants, irrespective of household reports of food (in)security (A-C, ANCOVA, p>0.05). Data are group median (IQR) for measured Apgar scores at one and five minutes postpartum. HUU = HIV-unexposed, uninfected infant; HEU = HIV-exposed, uninfected infant.

**Supplementary figure S5.**
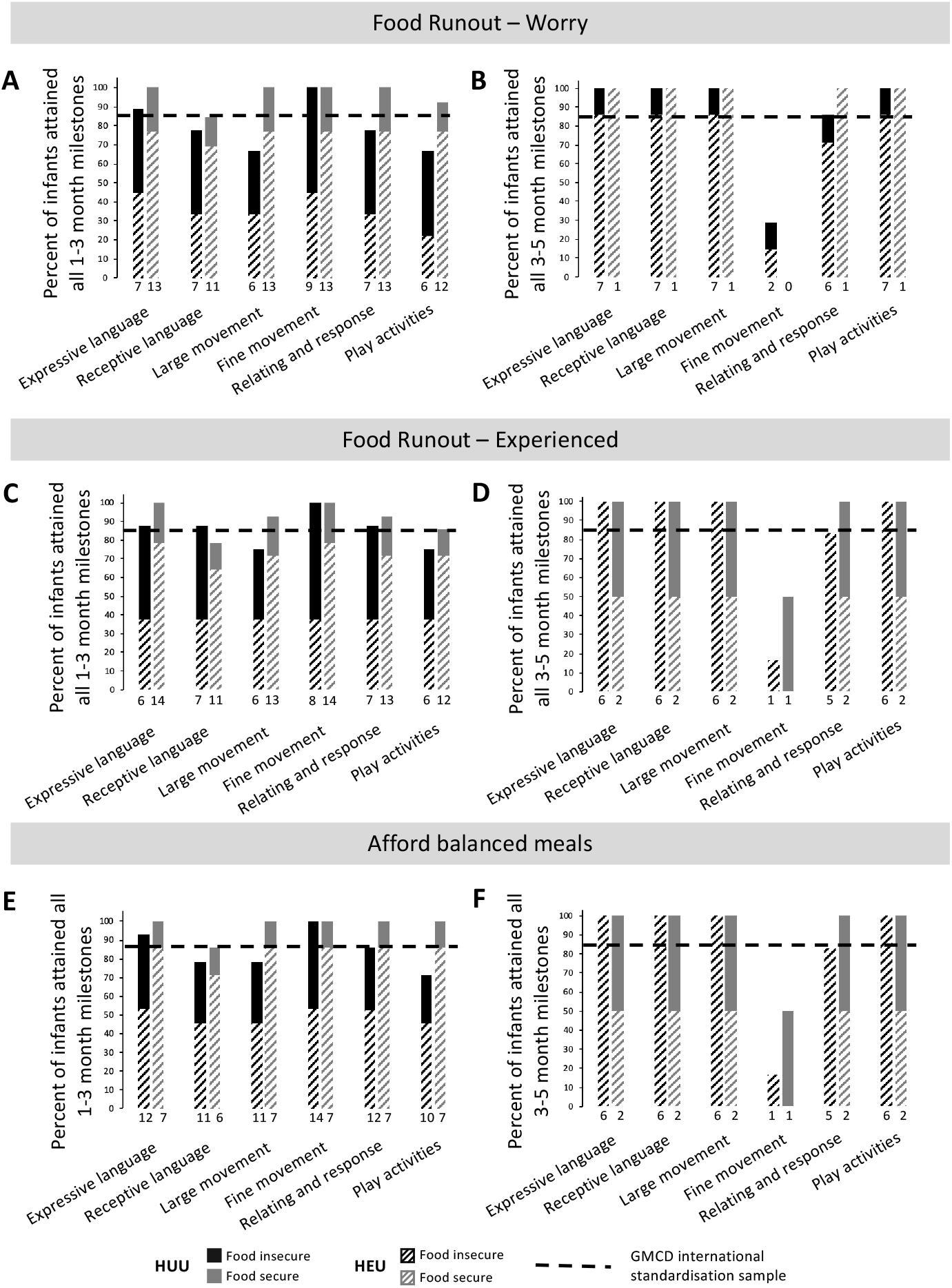
Food insecurity associates with low attainment of GMCD milestones for HUU and HEU infants. Infants whose mothers reported household food insecurity did not attain 1-3 month GMCD milestones (A, C, E) for receptive language, large movement, relating and response behaviour or play activities, or 3-5 month GMCD milestones (B, D, F) for fine movement or relating and response behaviour in the same proportion as the international standardization sample. Maternal reports of food insecurity did not associate with risk of not attaining all 1-3 month or 3-5 month GMCD milestones (A-F, [p>0.05], Fisher’s exact 2-Tail). Data are proportion (%) of infants who attained all age-appropriate GMCD milestones. The horizontal dotted line represents the GMCD standardised international sample proportion (85%) of infants who attained all milestones in that age category, when they were in that age range. The numbers underneath the bars represent the number of infants attaining all milestones for each milestone. GMCD = Guide for monitoring child development; HUU = HIV-unexposed, uninfected infant; HEU = HIV-exposed, uninfected infant.

**Supplementary table S1.**
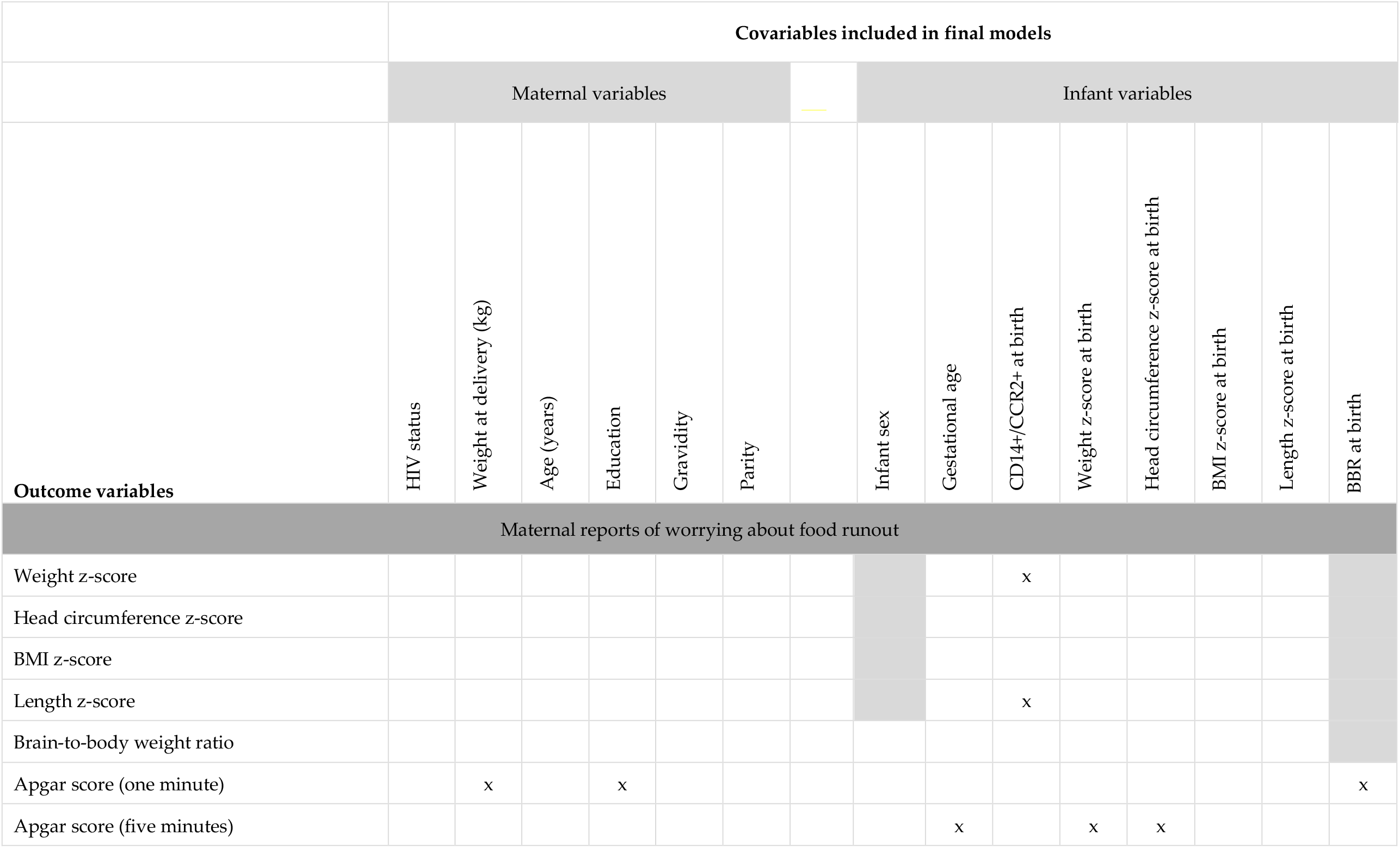
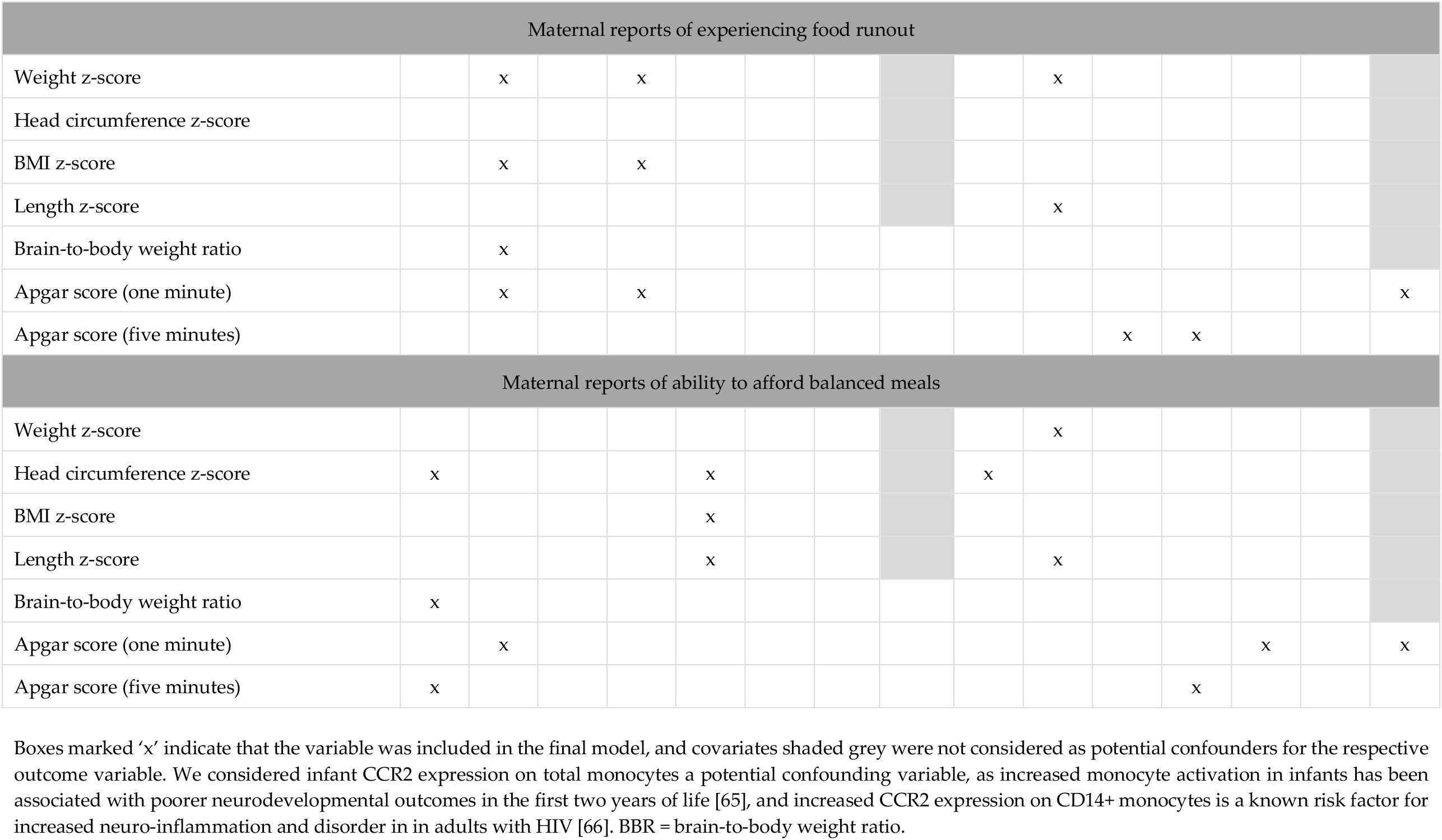
Final ANCOVA models for analysis of infant anthropometry and Apgar scores at birth for infants who were exposed to food insecure conditions compared to those who were not.

**Supplementary table S2.**
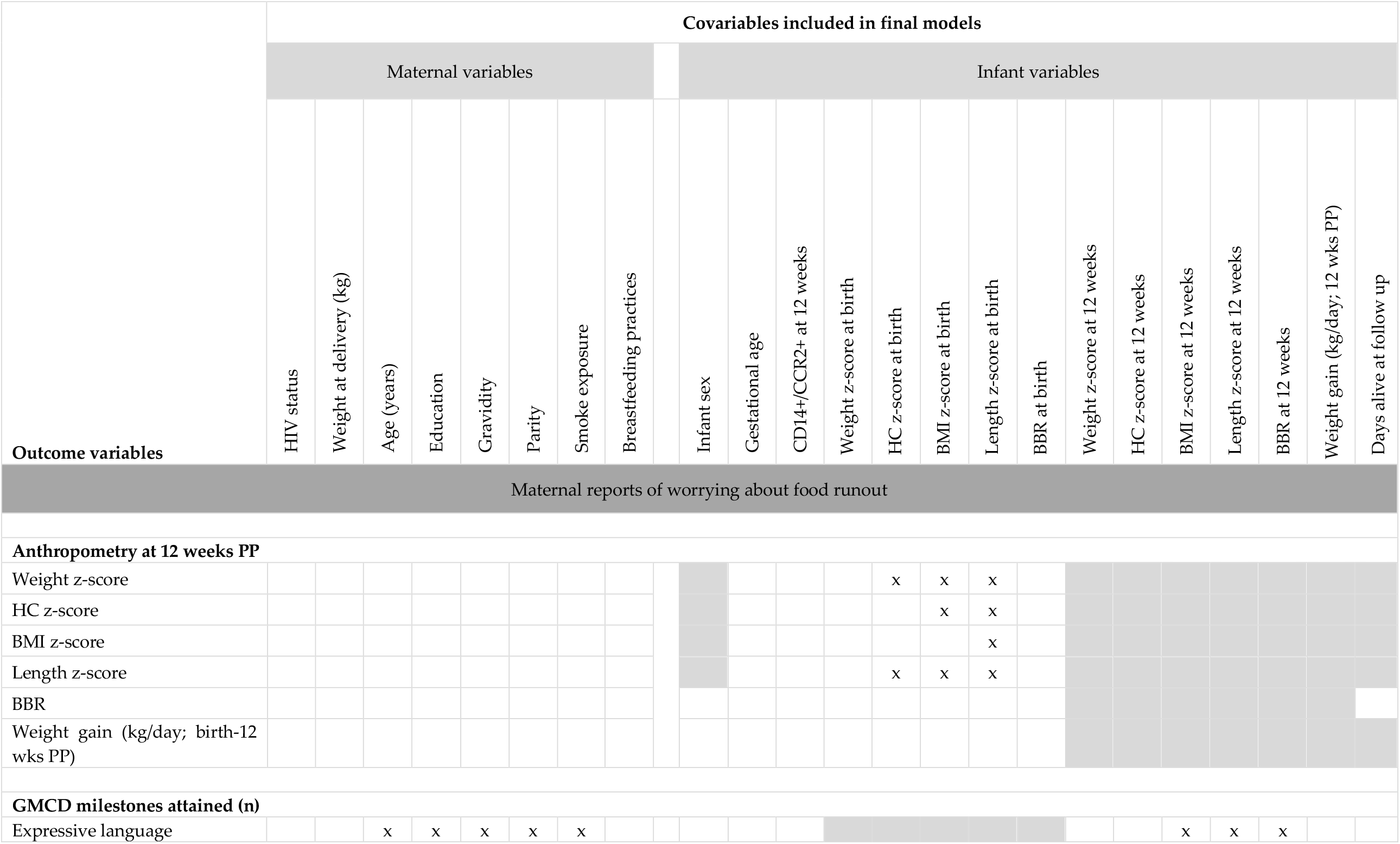
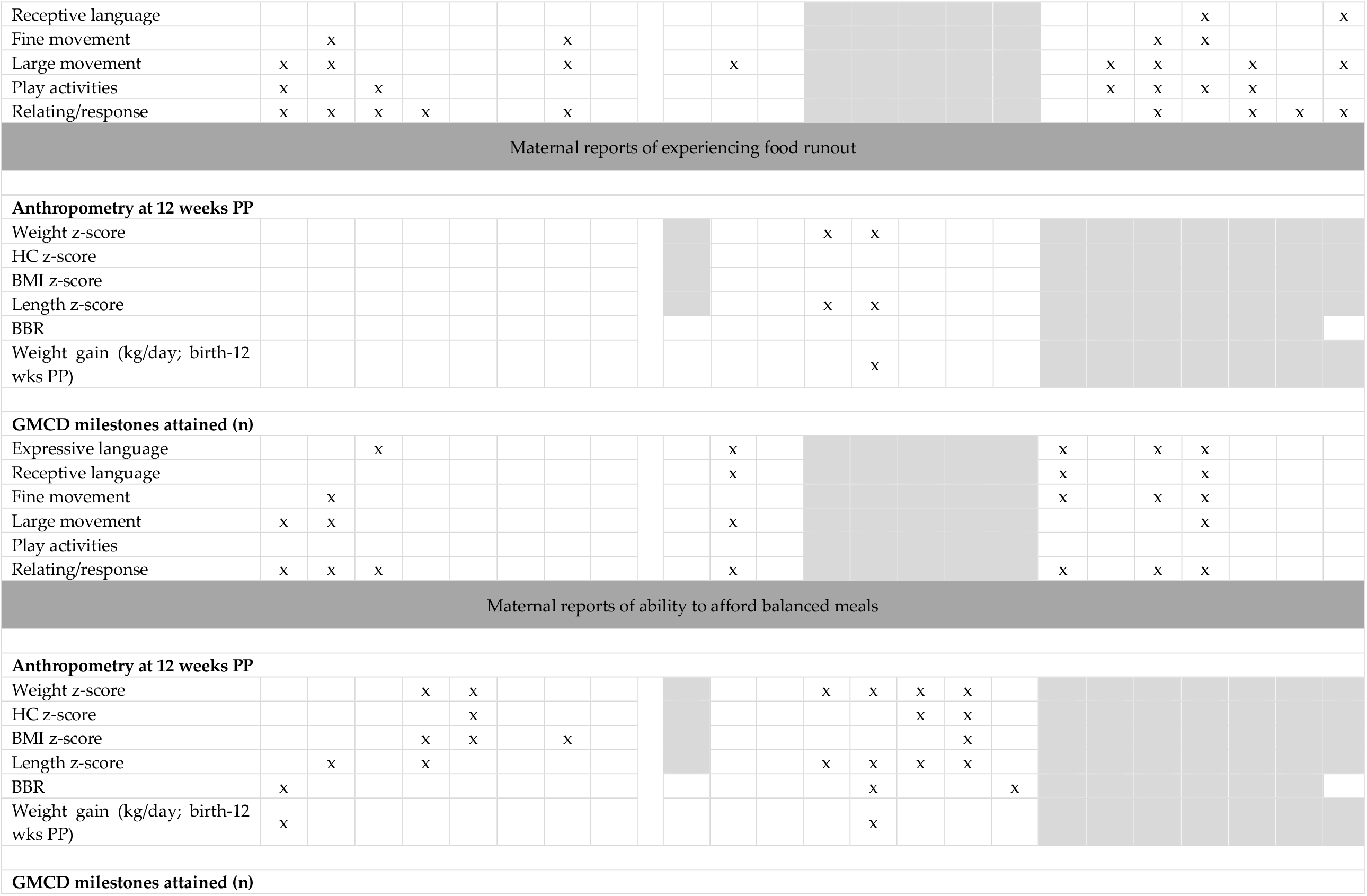
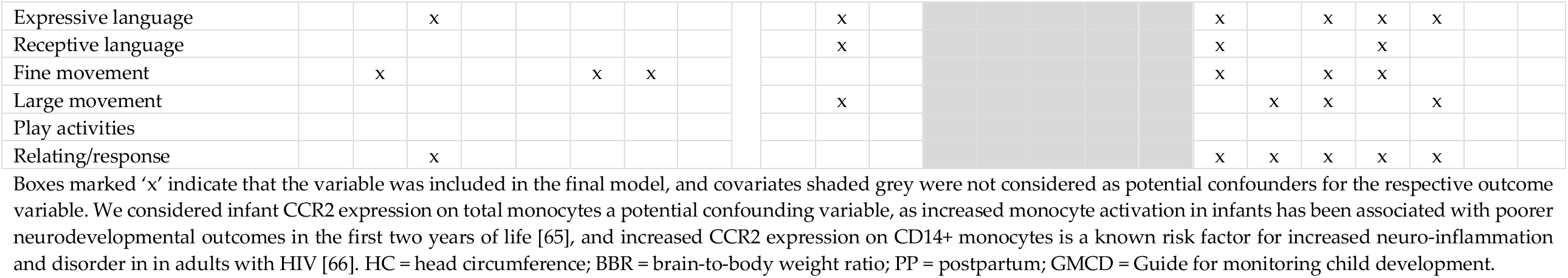
Final ANCOVA models for analysis of infant anthropometry and neurodevelopmental outcome at 12 weeks postpartum for infants who were exposed to food insecure conditions compared to those who were not.

**Supplementary Table S3.**
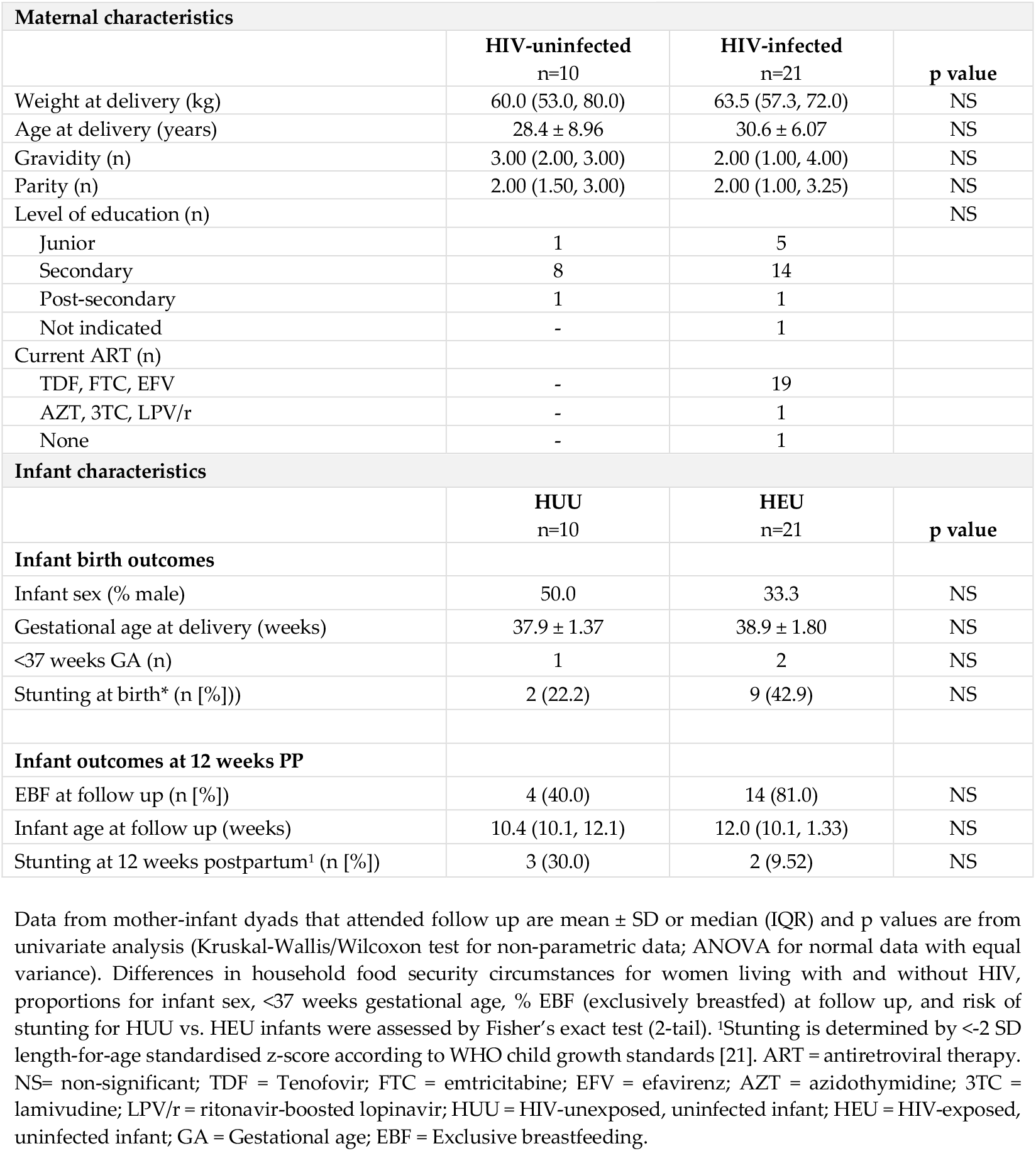
Maternal and infant cohort characteristics.

**Supplementary Table S4.** Anthropometry and Apgar scores at birth, and anthropometry, GMCD outcomes and breastfeeding practices at 12 weeks postpartum for infants exposed to food-insecure conditions compared to those who were not.

|  | Do you worry about food runout? |  |  | Do you experience food runout? |  |  | Are you able to afford balanced meals? |  |  |
| --- | --- | --- | --- | --- | --- | --- | --- | --- | --- |
|  | NO | YES | p value | NO | YES | p value | YES | NO | p value |
|  | n=14 | n=17 |  | n=16 | n=15 |  | n=9 | n=22 |  |
| **Gestational age at delivery (weeks)** | 38.8 ± 1.81 | 38.4 ± 1.66 | NS | 38.6 ± 1.79 | 38.5 ± 1.69 | NS | 38.2 ± 1.99 | 38.7 ± 1.62 | NS |
| **EBF at 12 weeks PP (n [%])** | 11 (78.6) | 7 (41.4) | NS | 13 (81.3) | 5 (33.3) | 0.01 | 8 (88.9) | 9 (42.9) | 0.04 |
| **Infant anthropometry and Apgar at birth\*** |  |  |  |  |  |  |  |  |  |
| Head circumference z-score | -1.49 ± 0.99 | -0.81 ± 1.46 | NS | -1.39 ± 0.96 | -0.82 ± 1.56 | NS | -1.71 ± 1.14 | -0.87 ± 1.30 | NS |
| Weight z-score | -0.59 ± 1.00 | -1.08 ± 1.10 | NS | -0.59 ± 0.94 | -1.14 ± 1.15 | NS | -0.83 ± 0.93 | -0.87 ± 1.14 | NS |
| Length z-score | -1.20 ± 1.41 | -1.05 ± 2.10 | NS | -1.21 ± 1.36 | -1.02 ± 2.20 | NS | -0.98 ± 1.35 | -1.18 ± 1.99 | NS |
| BMI z-score | -0.02 ± 1.81 | -0.94 ± 1.64 | NS | -0.02 ± 1.68 | -1.06 ± 1.69 | NS | -0.64 ± 1.61 | -0.50 ± 1.83 | NS |
| Brain-to-body weight ratio | 9.65 (8.90, 10.3) | 10.4 (9.70, 12.0) | 0.04 | 9.78 (8.95, 10.4) | 10.5 (9.54, 12.0) | NS | 9.54 (8.97, 10.4) | 10.1 (9.38, 11.4) | NS |
| Apgar 1-minute score | 9.00 (8.75, 9.00) | 8.00 (8.00, 9.00) | NS | 9.00 (8.25, 9.00) | 8.00 (8.00, 9.00) | NS | 9.00 (8.50, 9.00) | 8.00 (8.00, 9.00) | NS |
| Apgar 5-minute score | 10.0 (9.00, 10.0) | 9.00 (9.00, 9.00) | 0.02 | 9.50 (9.00, 10.0) | 9.00 (9.00, 9.00) | NS | 10.0 (9.00, 10.0) | 9.00 (9.00, 9.00) | NS |
| **Infant anthropometry in infants aged 12 weeks<sup>1</sup>** |  |  |  |  |  |  |  |  |  |
| Head circumference z-score | -1.47 (-2.12, -0.97) | -1.07 (-1.94, -0.62) | NS | -1.47 (-2.21, -0.88) | -1.07 (-1.94, -0.62) | NS | -1.47 (-2.17, -0.80) | -1.34 (-1.95, -0.79) | NS |
| Weight z-score | -0.08 (-0.77, 0.23) | -0.45 (-1.26, 0.14) | NS | -0.08 (-0.66, 0.26) | -0.46 (-1.47, 0.11) | NS | -0.41 (-0.83, 0.38) | -0.13 (-1.12, 0.14) | NS |
| Length z-score | -0.84 ± 0.64 | -1.52 ± 0.99 | NS | -0.85 ± 0.61 | -1.60 ± 1.02 | NS | -1.14 (-1.45, -0.69) | -1.02 (-1.80, -0.38) | NS |
| BMI z-score | 0.37 ± 0.96 | 0.19 ± 1.74 | NS | 0.41 ± 0.96 | 0.11 ± 1.82 | NS | 0.64 ± 1.00 | 0.12 ± 1.55 | NS |
| Brain-to-body weight ratio | 7.40 (6.84, 7.74) | 7.61 (7.11, 7.91) | NS | 7.21 (6.88, 7.69) | 7.72 (7.29, 8.12) | NS | 7.48 (7.05, 7.68) | 7.59 (7.03, 7.88) | NS |
| Weight gain (birth to 12 weeks PP; kg/day) | 0.03 ± 0.01 | 0.03 ± 0.01 | NS | 0.03 ± 0.01 | 0.03 ± 0.01 | NS | 0.03 ± 0.01 | 0.03 ± 0.01 | NS |
| **Total number of GMCD milestones attained in infants aged 12 weeks** |  |  |  |  |  |  |  |  |  |
| Fine movement | 1.00 (1.00, 1.00) | 1.00 (1.00, 1.00) | NS | 1.00 (1.00, 1.00) | 1.00 (1.00, 1.00) | NS | 1.00 (1.00, 1.50) | 1.00 (1.00, 1.00) | NS |
| Large movement | 4.00 (4.00, 4.00) | 4.00 (3.00, 4.00) | 0.03 | 4.00 (4.00, 4.00) | 4.00 (3.00, 4.00) | NS | 4.00 (4.00, 4.00) | 4.00 (3.00, 4.00) | NS |
| Expressive language | 3.00 (3.00, 3.00) | 3.00 (2.25, 3.00) | NS | 3.00 (3.00, 3.00) | 3.00 (2.75, 3.00) | NS | 3.00 (3.00, 3.00) | 3.00 (3.00, 3.00) | NS |
| Receptive language | 3.00 (3.00, 3.00) | 3.00 (3.00, 3.00) | NS | 3.00 (3.00, 3.00) | 3.00 (3.00, 3.00) | NS | 3.00 (3.00, 3.00) | 3.00 (3.00, 3.00) | NS |
| Play activities | 2.00 (2.00, 2.00) | 2.00 (1.00, 2.00) | 0.02 | 2.00 (2.00, 2.00) | 2.00 (1.00, 2.00) | NS | 2.00 (2.00, 2.00) | 2.00 (1.00, 2.00) | NS |
| Relating and response behaviour | 3.00 (2.75, 3.00) | 2.00 (2.00, 3.00) | NS | 3.00 (2.25, 3.00) | 2.00 (2.00, 3.00) | NS | 3.00 (3.00, 3.00) | 2.00 (2.00, 3.00) | NS |

Data from mother-infant dyads that attended follow up are presented mean ± SD or median (IQR) and p values are from adjusted multiple variable regression models. <sup>1</sup>All infant anthropometric measures are standardised according to WHO child growth standards [21]. Differences in proportion of infants exclusively breastfed (EBF) at follow up were assessed by Fisher's exact test (2-tail). PP = postpartum; GMCD = Guide for monitoring child development.

**Supplementary table S5.** Maternal nutrient intakes from one 24-hour dietary recall for mothers with and without HIV who attended follow up.

|  | **HIV-uninfected**<br>(n=10) | **HIV-infected**<br>(n=21) | **p value** |
| --- | --- | --- | --- |
| **Macronutrients – Absolute Intakes** |  |  |  |
| Energy (kJ) | 7339 (3862-7984) | 5204 (4024-6590) | NS |
| Nitrogen (g) | 4.15 (2.26-6.29) | 6.67 (4.87-8.20) | 0.02 |
| Total protein (g) | 46.6 (23.7-57.6) | 44.0 (33.2-61.6) | NS |
| Plant protein (g) | 26.0 (13.2-40.5) | 18.9 (12.5-25.4) | NS |
| Animal protein (g) | 13.8 (3.3-33.5) | 27.5 (18.6-35.0) | NS |
| Total fat (g) | 30.6 (22.8-81.4) | 33.9 (22.2-53.8) | NS |
| Carbohydrate, avail. (g) | 201 (140 - 282) | 160 (136-209) | NS |
| Starch (g) | 0.00 (0-4.55) | 5.50 (0.1-9.3) | 0.04 |
| Glucose (g) | 0.20 (0-5.8) | 3.70 (1.8-4.55) | NS |
| Fructose (g) | 0.20 (0-7.35) | 6.10 (0.65-6.8) | NS |
| Galactose (g) | 0.00 (0-0) | 0.00 (0-0) | - |
| Sucrose (g) | 10.2 (0-33.3) | 15.1 (11.8-22.1) | NS |
| Maltose (g) | 0.00 (0-0) | 0.00 (0-0) | - |
| Lactose (g) | 0.00 (0-3.38) | 0.00 (0-0.2) | NS |
| Total sugars (g) | 23.1 (6.83-33.8) | 29.0 (14.5-38.4) | NS |
| Added sugar (g) | 6.35 (0.53-67.0) | 13.4 (10.6-26.05) | NS |
| Non-starch polysaccharides (g) | 0.90 (0-10.5) | 3.60 (1.4-5.75) | NS |
| Insoluble NSP (g) | 0.55 (0-5.48) | 2.00 (0.9-2.8) | NS |
| Soluble NSP (g) | 0.35 (0-4.48) | 1.50 (0.55-2.6) | NS |
| Lignin (g) | 0.05 (0-0.38) | 0.20 (0.1-0.3) | NS |
| Total dietary fibre (g) | 17.0 (9.23-24.6) | 12.2 (8.85-15.8) | NS |
| Insoluble dietary fibre (g) | 0.60 (0-6.1) | 2.30 (0.95-3.2) | NS |
| Soluble dietary fibre (g) | 0.35 (0-4.48) | 1.50 (0.55-2.6) | NS |
| Ash (g) | 7.75 (3.8-13.8) | 6.10 (4.6-12.5) | NS |
| Moisture (g) | 623 (340-1073) | 789 (628-1030) | NS |
| **Minerals – Absolute Intakes** |  |  |  |
| Ca (mg) | 195 (106-382) | 132 (85-187) | NS |
| Fe (mg) | 11.3 (6.98-13.5) | 10.9 (7.4-12.5) | NS |
| Haem iron (mg) | 0.05 (0-0.63) | 0.40 (0-0.55) | NS |
| Non-haem iron (mg) | 1.95 (0.15-4.55) | 3.30 (2.25-4.25) | NS |
| Mg (mg) | 280 (116-347) | 168 (135-224) | NS |
| P (mg) | 723 (418-855) | 573 (420-731) | NS |
| K (mg) | 1838 (910-2858) | 1217 (925-1474) | NS |
| Na (mg) | 2126 (1009-3923) | 1608 (1010-3360) | NS |
| Cl (mg) | 96.5 (30-406) | 397 (245-567) | 0.02 |
| Zn (mg) | 6.95 (5.16-9.53) | 9.46 (7.81-11.97) | NS |
| Cu (mg) | 1.11 (0.48-1.27) | 0.74 (0.5-0.96) | NS |
| Cr (mcg) | 10.4 (0-47.03) | 31.00 (25.3-49.8) | 0.05 |
| Se (mcg) | 7.45 (4.38-20.3) | 30.5 (24.9-43.5) | <.001 |
| Mn (mcg) | 1730 (716-2414) | 991 (801-1782) | NS |
| I (mcg) | 91.0 (25.3-168) | 92.0 (63.5-189) | NS |
| B (mcg) | 71.5 (0-11) | 438 (169-624) | NS |
| F (mcg) | 63.5 (0.75-153) | 111 (93-141) | NS |
| Si (mcg) | 0.00 (0-842) | 1776 (284-3838) | 0.01 |
| **Vitamins – Absolute Intakes** |  |  |  |
| Vitamin A (RE) (mcg) | 423 (330-690) | 430 (301-705) | NS |
| Retinol (mcg) | 1.50 (0-73.75) | 60.0 (33-75.5) | NS |
| Total carotenoids (mcg) | 745 (0-3408) | 384 (162-1434) | NS |
| β-Carotene (mcg) | 627 (0-3368) | 281 (156-1265) | NS |
| α-Carotene (mcg) | 0.00 (0-36.3) | 104 (0-123) | NS |
| Cryptoxanthin (mcg) | 5.00 (0-39.3) | 16.0 (1-23.5) | NS |
| Thiamin (mg) | 1.06 (0.64-1.40) | 1.08 (0.69-1.41) | NS |
| Riboflavin (mg) | 0.51 (0.32-1.26) | 0.76 (0.65-0.97) | NS |
| Niacin (mg) | 14.6 (9.03-20.28) | 16.3 (9.8-19.6) | NS |
| Vitamin B6 (mg) | 2.36 (1.37-5.19) | 3.13 (1.66-3.723) | NS |
| Folate (mcg) | 212 (180-383) | 278 (207-312) | NS |
| Vitamin B12 (mcg) | 1.65 (0.23-2.45) | 1.80 (1.6-3.2) | NS |
| Pantothenate (mg) | 2.33 (2.08-3.02) | 2.95 (2.27-4.82) | NS |
| Biotin (mcg) | 14.5 (10.1-27.8) | 25.4 (20.8-34.7) | NS |
| Vitamin C (mg) | 31.5 (2.25-91.5) | 22.0 (10.5-62.5) | NS |
| Vitamin D (mcg) | 0.86 (0.04-2.08) | 6.45 (4.20-8.46) | 0.002 |
| Vitamin E (mg) | 5.24 (3.32-11.7) | 7.91 (4.73-13.3) | NS |
| α-Tocopherol (mg) | 0.38 (0-3.72) | 6.08 (3.44-8.67) | 0.005 |
| β-Tocopherol (mg) | 0.00 (0-0.11) | 0.20 (0.12-0.31) | 0.002 |
| δ-Tocopherol (mg) | 0.00 (0-0.02) | 0.11 (0.11-0.22) | <.001 |
| γ-Tocopherol (mg) | 0.00 (0-0.21) | 0.49 (0.45-0.83) | 0.004 |
| α-Tocotrienol (mg) | 0.00 (0-0.06) | 0.07 (0.07-0.15) | 0.005 |
| β-Tocotrienol (mg) | 0.00 (0-0.02) | 0.05 (0.03-0.06) | 0.005 |
| δ-Tocotrienol (mg) | 0.00 (0-0) | 0.00 (0-0) | - |
| γ-Tocotrienol (mg) | 0.00 (0-0.02) | 0.01 (0-0.02) | NS |
| Lycopene (mcg) | 0.00 (0-133) | 0.00 (0-2.5) | NS |
| Lutein (mcg) | 38.0 (0-7900) | 51.0 (9-140) | NS |
| Vitamin K (mcg) | 23.8 (0.05-403) | 19.8 (8.9-57.2) | NS |
| **Fatty Acids (FA) and Cholesterol – Absolute Intakes** |  |  |  |
| Saturated FA (g) | 9.31 (5.67-14.9) | 8.95 (6.34-13.2) | NS |
| Mono-unsaturated FA (g) | 11.8 (7.01-19.6) | 11.8 (7.90-16.4) | NS |
| Polyunsaturated FA (g) | 6.74 (2.74-37.4) | 7.15 (5.36-16.0) | NS |
| Single trans FA (g) | 0.00 (0-0) | 0.00 (0-0) | - |
| Double trans FA (g) | 0.00 (0-0) | 0.00 (0-0) | - |
| Total trans FA (g) | 0.13 (0-1.27) | 0.19 (0.03-0.34) | NS |
| Cholesterol (mg) | 65.5 (30-130) | 324 (259-461) | <.0001 |
| C4:0 (g) | 0.00 (0-0.11) | 0.00 (0-0) | NS |
| C6:0 (g) | 0.00 (0-0.07) | 0.00 (0-0) | NS |
| C8:0 (g) | 0.00 (0-0.04) | 0.00 (0-0.02) | NS |
| C10:0 (g) | 0.01 (0-0.09) | 0.02 (0-0.05) | NS |
| C12:0 (g) | 0.04 (0.01-0.21) | 0.04 (0.01-0.13) | NS |
| C13:0 (g) | 0.00 (0-0) | 0.00 (0-0) | - |
| C14:0 (g) | 0.36 (0.06-0.94) | 0.27 (0.18-0.45) | NS |
| C15:0 (g) | 0.00 (0-0) | 0.02 (0.01-0.02) | <.001 |
| C16:0 (g) | 5.10 (3.04 - 8.57) | 4.81 (3.68-7.16) | NS |
| C17:0 (g) | 0.00 (0-0.01) | 0.01 (0-0.01) | NS |
| C18:0 (g) | 2.25 (1.36-3.23) | 2.25 (1.9-3.72) | NS |
| C20:0 (g) | 0.09 (0.02-0.20) | 0.05 (0.02-0.1) | NS |
| C21:0 (g) | 0.00 (0-0) | 0.00 (0-0) | - |
| C22:0 (g) | 0.05 (0-0.25) | 0.04 (0.02-0.15) | NS |
| C23:0 (g) | 0.00 (0-0) | 0.00 (0-0) | - |
| C24:0 (g) | 0.02 (0-0.10) | 0.04 (0.02-0.10) | NS |
| C10:1 (g) | 0.00 (0-0) | 0.00 (0-0) | - |
| C12:1 (g) | 0.00 (0-0) | 0.00 (0-0) | - |
| C14:1 (g) | 0.00 (0-0.04) | 0.01 (0-0.02) | NS |
| C15:1 (g) | 0.00 (0-0) | 0.00 (0-0) | - |
| C16:1 (g) | 0.33 (0.19-0.53) | 0.60 (0.48-0.97) | 0.01 |
| C17:1 (g) | 0.00 (0-0) | 0.00 (0-0) | NS |
| C18:1 (g) | 10.49 (5.57-18.4) | 10.09 (6.54-14.5) | NS |
| C20:1 (g) | 0.10 (0.03-0.25) | 0.04 (0.02-0.11) | NS |
| C22:1 (g) | 0.04 (0-0.21) | 0.00 (0-0.03) | NS |
| C23:1 (g) | 0.00 (0-0) | 0.00 (0-0) | - |
| C24:1 (g) | 0.00 (0-0) | 0.00 (0-0) | - |
| C18:2 (g) | 5.71 (1.42-35.7) | 5.54 (4.09-12.3) | NS |
| C18:3 (g) | 0.26 (0.11-0.99) | 0.21 (0.14-0.4) | NS |
| C18:4 (g) | 0.00 (0-0.005) | 0.00 (0-0) | NS |
| C20:2 (g) | 0.00 (0-0.01) | 0.04 (0.03-0.06) | 0.001 |
| C20:4 (g) | 0.04 (0-0.07) | 0.07 (0.04-0.1) | NS |
| C20:5 (g) | 0.02 (0-0.04) | 0.03 (0.02-0.04) | NS |
| C22:2 (g) | 0.00 (0-0.003) | 0.01 (0-0.01) | 0.03 |
| C22:3 (g) | 0.00 (0-0) | 0.00 (0-0) | - |
| C22:4 (g) | 0.00 (0-0) | 0.00 (0-0) | - |
| C22:5 (g) | 0.00 (0-0.01) | 0.00 (0-0.01) | NS |
| C22:6 (g) | 0.03 (0-0.07) | 0.07 (0.04-0.08) | NS |
| C24:6 (g) | 0.00 (0-0) | 0.00 (0-0) | - |
| C20:3 (g) | 0.00 (0-0.01) | 0.00 (0-0.01) | NS |

| Amino Acids – Absolute Intakes |  |  |  |
| --- | --- | --- | --- |
| Isoleucine (g) | 1.46 (0.74-2.10) | 1.41 (1.16-2.16) | NS |
| Leucine (g) | 2.76 (1.38-4.08) | 2.78 (2.14-3.88) | NS |
| Lysine (g) | 1.72 (0.98-2.90) | 2.46 (1.55-3.195) | NS |
| Methionine (g) | 0.83 (0.41-0.96) | 0.92 (0.65-1.19) | NS |
| Phenylalanine (g) | 1.59 (0.79-2.39) | 1.59 (1.21-2.36) | NS |
| Threonine (g) | 1.31 (0.70-1.92) | 1.43 (1.07-1.98) | NS |
| Tryptophan (g) | 0.46 (0.27-0.63) | 0.54 (0.38-0.71) | NS |
| Valine (g) | 1.72 (0.85-2.47) | 1.68 (1.36-2.49) | NS |
| Arginine (g) | 1.69 (1.13-2.83) | 2.23 (1.53-3.27) | NS |
| Histidine (g) | 0.88 (0.41-1.24) | 0.93 (0.73-1.29) | NS |
| Cystine (g) | 0.33 (0.20-0.48) | 0.64 (0.53-0.81) | <.001 |
| Tyrosine (g) | 0.69 (0.29-1.02) | 1.06 (0.73-1.40) | 0.03 |
| Alanine (g) | 0.85 (0.35-1.60) | 1.59 (1.11-2.23) | 0.02 |
| Aspartic acid (g) | 1.45 (0.80-2.69) | 2.90 (1.78-3.53) | 0.04 |
| Glutamic acid (g) | 4.76 (3.04-5.91) | 5.50 (3.31-8.31) | NS |
| Glycine (g) | 0.73 (0.36-1.44) | 1.33 (0.93-2.00) | 0.03 |
| Proline (g) | 1.43 (0.89-2.01) | 1.74 (1.09-2.58) | NS |
| Serine (g) | 0.98 (0.46-1.59) | 1.65 (1.33-2.02) | 0.01 |
| Hydroxyproline (g) | 0.02 (0-0.18) | 0.09 (0-0.17) | NS |
| Alcohol (g) | 0.00 (0-0) | 0.00 (0-0) | - |
| Phytate (mg) | 155 (6-192) | 192 (99.5-225) | NS |
| Malic acid (mg) | 38.0 (0-879) | 305 (14.5-612.5) | NS |
| Citric acid (mg) | 276.50 (0-1275) | 221 (77-508) | NS |
| Tartaric acid (mg) | 0.00 (0-0) | 0.00 (0-0) | NS |
| Oxalic acid (mg) | 6.00 (0-384) | 11.0 (0-37.5) | NS |
| Caffeine (mg) | 0.00 (0-0) | 0.00 (0-0) | - |
| Tannins (mg) | 0.00 (0-0) | 0.00 (0-0) | - |
| Energy – % |  |  |  |
| % Energy - Protein | 0.13 (0.08-0.18) | 0.15 (0.13-0.18) | NS |
| % Energy - Fat | 0.20 (0.13-0.40) | 0.25 (0.21-0.31) | NS |
| % Energy - Saturated SFA | 0.06 (0.04-0.07) | 0.06 (0.05-0.09) | NS |
| % Energy - Mono-unsaturated FA | 0.07 (0.05-0.12) | 0.09 (0.08-0.11) | NS |
| % Energy – Polyunsaturated FA | 0.04 (0.02-0.16) | 0.06 (0.04-0.09) | NS |
| % Energy - Carbohydrate | 0.64 (0.52-0.70) | 0.59 (0.51-0.65) | NS |

Data from mother-infant dyads that attended follow up are presented as median (IQR) and p values are from univariate analysis (Kruskal-Wallis/Wilcoxon test for non-parametric data; ANOVA for normal data with equal variance). Iodised salt was assumed to be consumed by all mothers, as the majority of salt consumed in South Africa is iodised [64].

**Supplementary table S6.** Maternal nutrient intake from one 24-hour dietary recall for mothers who report experiencing food insecurity compared to those who do not experience food insecurity.

|  | Do you experience food runout? |  |  | Are you able to afford balanced meals? |  |  |
| --- | --- | --- | --- | --- | --- | --- |
|  | NO | YES | p value | YES | NO | p value |
|  | n=16 | n=15 |  | n=9 | n=22 |  |
| **Dietary diversity score (/9)** | 5.00 (4.00, 6.00) | 3.00 (4.00, 5.00) | NS | 4.00 (3.50, 5.50) | 4.00 (3.75, 6.00) | NS |
| **Dietary diversity score <4 (n)** | 2 | 5 | NS | 2 | 5 | NS |
| **Absolute intakes** |  |  |  |  |  |  |
| Macronutrients |  |  |  |  |  |  |
| Moisture (g) | 868 (620, 1030) | 670 (409, 1011) | NS | 789 (614, 1037) | 806 (564, 1016) | NS |
| Energy (kJ) | 5601 (4659, 7004) | 5204 (3247, 8339) | NS | 5691 (4519, 7046) | 5358 (3538, 7505) | NS |
| Nitrogen (g) | 7.15 (5.68, 9.29) | 4.31 (3.87, 6.10) | 0.02 | 6.94 (5.00, 9.53) | 5.52 (4.03, 7.37) | NS |
| Total protein (g) | 51.4 (42.5, 68.7) | 32.9 (26.1, 44.7) | 0.03 | 53.3 (41.5, 67.7) | 40.9 (27.5, 54.6) | NS |
| Plant protein (g) | 19.6 (15.8, 27.3) | 22.3 (11.5, 34.9) | NS | 19.9 (17.0, 33.1) | 20.6 (11.6, 34.0) | NS |
| Animal protein (g) | 29.0 (19.7, 38.0) | 15.4 (3.30, 21.4) | 0.01 | 28.5 (19.0, 38.5) | 18.9 (9.65, 33.2) | NS |
| Total fat (g) | 35.0 (29.9, 45.8) | 25.8 (19.6, 69.0) | NS | 37.5 (25.1, 57.2) | 31.0 (21.9, 63.1) | NS |
| Carbohydrates, avail. (g) | 162 (159, 234) | 161 (93.8, 280) | NS | 165 (158, 229) | 160 (106, 259) | NS |
| Starch (g) | 5.60 (0.13, 10.1) | 0.00 (0.00, 5.40) | NS | 5.60 (0.05, 6.80) | 0.20 (0.00, 12.5) | NS |
| Glucose (g) | 3.95 (2.75, 5.60) | 1.20 (0.00, 3.40) | 0.01 | 3.80 (2.50, 5.25) | 2.55 (0.30, 4.55) | NS |
| Fructose (g) | 6.50 (1.20, 8.15) | 0.40 (0.00, 6.10) | 0.01 | 6.50 (1.90, 7.75) | 1.10 (0.00, 6.65) | NS |
| Sucrose (g) | 19.1 (12.4, 30.1) | 8.40 (1.00, 17.9) | 0.03 | 20.4 (16.3, 28.1) | 12.0 (3.98, 23.9) | NS |
| Maltose (g)<sup>1</sup> |  | 0.1 |  |  | 0.1 |  |
| Galactose<sup>2</sup> |  |  |  |  |  |  |
| Lactose (g) | 0.00 (0.00, 1.60) | 0.00 (0.00, 0.10) | NS | 0.00 (0.00, 2.40) | 0.00 (0.00, 0.15) | NS |
| Total sugars (g) | 31.2 (24.3, 42.5) | 13.1 (7.00, 30.9) | 0.02 | 32.0 (29.6, 39.5) | 20.8 (9.10, 37.1) | NS |
| Added sugar (g) | 17.6 (12.0, 40.5) | 8.00 (0.30, 21.0) | 0.04 | 13.4 (10.5, 25.6) | 12.0 (3.70, 39.5) | NS |
| Total dietary fibre (g) | 13.4 (10.9, 17.2) | 12.2 (6.20, 18.4) | NS | 14.5 (11.6, 16.3) | 12.3 (8.10, 17.9) | NS |
| Insoluble dietary fibre (g) | 3.15 (1.68, 4.08) | 1.20 (0.00, 3.20) | 0.04 | 3.10 (1.35, 3.90) | 1.75 (0.00, 3.33) | NS |
| Soluble dietary fibre (g) | 2.20 (1.30, 2.70) | 0.70 (0.00, 2.90) | NS | 2.20 (0.85, 2.75) | 1.30 (0.00, 2.75) | NS |
| Ash (g) | 6.05 (4.55, 6.58) | 10.5 (4.20, 14.8) | NS | 6.10 (4.25, 6.55) | 6.15 (4.63, 14.6) | NS |
| Non-starch polysaccharides (g) | 4.85 (2.68, 6.33) | 1.80 (0.00, 5.90) | NS | 4.90 (2.00, 6.25) | 2.75 (0.00, 6.05) | NS |
| Insoluble NSP (g) | 2.60 (1.43, 3.50) | 1.10 (0.00, 2.80) | NS | 2.50 (1.05, 3.30) | 1.45 (0.00, 2.98) | NS |
| Soluble NSP (g) | 2.20 (1.30, 2.70) | 0.70 (0.00, 2.90) | NS | 2.20 (0.85, 2.75) | 0.00 (1.30, 2.75) | NS |
| Lignin (g) | 0.25 (0.13, 0.38) | 0.00 (0.00, 0.20) | 0.01 | 0.30 (0.20, 0.40) | 0.10 (0.00, 0.23) | NS |
| Amino acids |  |  |  |  |  |  |
| Isoleucine (g) | 1.78 (1.38, 2.43) | 1.08 (0.79, 1.47) | 0.05 | 1.77 (1.26, 2.55) | 1.50 (0.89, 1.95) | NS |
| Leucine (g) | 3.48 (2.77, 4.47) | 1.95 (1.47, 2.75) | 0.03 | 3.36 (2.60, 2.46) | 2.47 (1.58, 3.71) | NS |
| lysine (g) | 2.59 (1.99, 3.80) | 1.23 (1.03, 2.57) | 0.03 | 2.71 (1.81, 4.05) | 1.74 (1.06, 2.65) | NS |
| Methionine (g) | 0.96 (0.88, 1.13) | 0.61 (0.45, 0.85) | 0.002 | 1.06 (0.93, 2.44) | 1.01 (0.78, 1.64) | NS |
| Phenylalanine (g) | 2.01 (1.55, 2.51) | 1.22 (0.80, 1.69) | 0.04 | 1.96 (1.42, 2.43) | 1.51 (1.00, 2.22) | NS |
| Threonine (g) | 1.73 (1.38, 2.22) | 0.95 (0.74, 1.43) | 0.02 | 1.69 (1.26, 2.23) | 1.25 (0.79, 1.81) | NS |
| Tryptophan (g) | 0.61 (0.53, 0.76) | 0.34 (0.30, 0.51) | 0.003 | 0.59 (0.48, 0.77) | 0.42 (0.31, 0.63) | NS |
| Valine (g) | 2.12 (1.68, 2.56) | 1.24 (0.87, 1.75) | 0.04 | 2.01 (1.53, 2.58) | 1.57 (1.12, 2.38) | NS |
| Arginine (g) | 2.53 (1.96, 3.57) | 1.48 (1.17, 2.12) | 0.03 | 2.44 (1.79, 3.49) | 1.80 (1.28, 2.68) | NS |
| Histidine (g) | 1.14 (0.89, 1.51) | 0.64 (0.42, 1.26) | 0.01 | 1.16 (0.82, 1.51) | 0.76 (0.50, 1.20) | NS |
| Cystine (g) | 0.62 (0.52, 0.81) | 0.48 (0.30, 0.62) | 0.02 | 0.60 (0.48, 0.78) | 0.52 (0.34, 0.72) | NS |
| Tyrosine (g) | 1.14 (0.94, 1.42) | 0.75 (0.46, 0.90) | 0.03 | 1.09 (0.80, 1.40) | 0.85 (0.49, 1.10) | NS |
| Alanine (g) | 1.75 (1.37, 2.29) | 0.92 (0.66, 1.46) | 0.006 | 1.76 (1.20, 2.32) | 1.24 (0.73, 1.73) | NS |
| Aspartic acid (g) | 3.01 (2.46, 3.74) | 1.47 (1.04, 2.90) | 0.04 | 2.93 (2.06, 3.90) | 1.87 (1.30, 3.02) | NS |
| Glutamic acid (g) | 5.85 (5.25, 8.12) | 4.33 (1.70, 5.78) | NS | 5.71 (4.26, 7.54) | 4.58 (2.39, 6.46) | NS |
| Glycine (g) | 1.52 (1.13, 2.14) | 0.84 (0.42, 1.30) | 0.009 | 1.49 (1.06, 2.13) | 1.04 (0.63, 1.48) | NS |
| Proline (g) | 1.85 (1.58, 2.59) | 1.28 (0.51, 1.92) | 0.03 | 1.81 (1.36, 2.35) | 1.38 (0.78, 2.36) | NS |
| Serine (g) | 1.68 (1.52, 2.02) | 1.09 (0.85, 1.48) | 0.01 | 1.65 (1.29, 2.02) | 1.43 (0.91, 1.71) | NS |
| Hydroxyproline (g) | 0.11 (0.06, 0.26) | 0.00 (0.00, 0.07) | 0.005 | 0.11 (0.07, 0.24) | 0.00 (0.00, 0.11) | 0.04 |
| Fatty acids and cholesterol |  |  |  |  |  |  |
| Saturated FA (g) | 9.68 (8.88, 14.7) | 6.80 (5.69, 12.9) | 0.03 | 9.77 (8.33, 19.5) | 7.96 (5.98, 12.1) | NS |
| Mono-unsaturated FA (g) | 11.7 (10.4, 15.0) | 12.0 (6.86, 25.7) | NS | 11.8 (8.26, 16.4) | 11.5 (7.6, 21.8) | NS |
| Polyunsaturated FA (g) | 7.67 (5.87, 13.9) | 5.47 (3.88, 22.7) | NS | 9.40 (4.85, 15.5) | 6.34 (4.49, 18.4) | NS |
| Single trans FA (g)<sup>2</sup> |  |  |  |  |  |  |
| Double trans FA (g)<sup>2</sup> |  |  |  |  |  |  |
| Total trans FA (g) | 0.21 (0.16, 0.75) | 0.04 (0.00, 0.20) | 0.03 | 0.73 (0.16, 1.00) | 0.06 (0.00, 0.21) | 0.03 |
| Cholesterol (mg) | 291 (191, 524) | 112 (26.0, 416) | 0.03 | 269 (213, 326) | 261 (45.5, 446) | NS |
| **Median intake (%) of TULs by mothers** |  |  |  |  |  |  |
| **Minerals** |  |  |  |  |  |  |
| Calcium (Ca) | 5.88 (4.56, 9.90) | 4.52 (2.72, 9.56) | NS | 7.04 (4.90, 11.7) | 4.76 (3.01, 8.33) | NS |
| Iron (Fe) | 25.6 (21.0, 27.4) | 18.2 (12.0, 30.2) | NS | 25.8 (21.3, 30.0) | 24.2 (14.6, 28.3) | NS |
| Magnesium (Mg) | 52.1 (43.1, 74.6) | 57.1 (30.3, 95.1) | NS | 54.0 (46.4, 78.7) | 52.1 (30.7, 88.6) | NS |
| Phosphorus (P) | 16.7 (12.8, 20.7) | 12.1 (8.65, 18.5) | NS | 15.2 (12.9, 21.2) | 14.6 (9.83, 18.9) | NS |
| Sodium (Na) | 61.9 (47.6, 84.9) | 132 (51.2, 221) | NS | 65.1 (48.4, 95.0) | 73.6 (45.0, 181) | NS |
| Zinc (Zn) | 23.3 (20.2, 30.3) | 17.7 (14.4, 24.2) | NS | 20.9 (20.2, 36.8) | 21.0 (15.2, 26.6) | NS |
| Copper (Cu) | 8.15 (6.43, 11.7) | 6.90 (3.90, 11.0) | NS | 8.60 (6.30, 10.7) | 7.15 (4.45, 11.4) | NS |
| Selenium (Se) | 7.63 (5.91, 11.2) | 5.58 (1.80, 7.78) | NS | 7.50 (6.03, 8.66) | 6.45 (2.73, 9.89) | NS |
| Manganese (Mn) | 12.8 (8.77, 19.3) | 12.7 (5.36, 16.7) | NS | 14.7 (8.84, 27.1) | 11.5 (6.52, 16.9) | NS |
| Iodine (I) | 7.67 (3.73, 11.2) | 14.0 (5.09, 19.6) | NS | 8.18 (3.59, 13.6) | 8.36 (4.84, 18.4) | NS |
| **Vitamins** |  |  |  |  |  |  |
| Vitamin A | 14.2 (10.5, 20.9) | 14.1 (8.54, 25.2) | NS | 12.2 (10.2, 16.1) | 14.7 (8.95, 24.9) | NS |
| Niacin | 47.4 (41.4, 55.9) | 40.0 (23.4, 55.7) | NS | 46.6 (41.7, 54.0) | 42.1 (24.9, 58.1) | NS |
| Vitamin B6 | 3.20 (2.72, 3.89) | 1.89 (1.16, 3.80) | NS | 3.26 (2.80, 5.09) | 2.61 (1.37, 3.69) | NS |
| Folate | 27.8 (20.0, 30.1) | 24.1 (20.5, 47.6) | NS | 27.8 (22.6, 29.9) | 25.0 (19.3, 37.1) | NS |
| Vitamin C | 1.34 (0.99, 2.55) | 0.55 (0.00, 6.25) | NS | 2.40 (0.85, 3.90) | 1.13 (0.23, 3.59) | NS |
| Vitamin D | 4.73 (4.14, 8.60) | 1.39 (0.05, 7.86) | NS | 4.72 (4.20, 6.22) | 4.12 (0.72, 8.06) | NS |
| Vitamin E | 0.73 (0.49, 1.16) | 0.68 (0.39, 1.46) | NS | 0.80 (0.51, 1.13) | 0.62 (0.42, 1.50) | NS |

Data from mother-infant dyads that attended follow up are presented as mean ± SD or median (IQR) and p values are from univariate analysis (Kruskal-Wallis/Wilcoxon test for non-parametric data; ANOVA for normal data with equal variance). Dietary diversity score was calculated using nine food groups, only counting each food groups once (refs). Percent intake of TULs were calculated using the Institute of Medicine's TULs for minerals and vitamins for
lactating women 14-18, 19-30 or 31-50 years of age [26]. Iodised salt was assumed to be consumed by all mothers, as the majority of salt consumed in South Africa is iodised [64]. TULs = Tolerable upper levels. <sup>1</sup>Only one mother consumed maltose. <sup>2</sup>None consumed by mothers.

## Footnotes

† Cited in Table 1 legend

‡ Cited in Supplementary Table S1 and S2 legends

